# Multi-Agent Generative Artificial Intelligence for Transfusion Reaction Feature Extraction

**DOI:** 10.64898/2026.11.06.26367389

**Authors:** Ayushi Kumari, Jeremy W. Jacobs, Laura Dilly, Aman Anand, Jennifer Woo, Yulia Lin, Andrew W. Shih, Garrett S. Booth, Ismail Raslan, Nicole Relke, Rounak Dubey, Siddhartha Srivastava, Widad Abdulwahab, Ruchika Goel, Sheharyar Raza

**Author notes:** **Correspondence: Sheharyar Raza**, MD Sunnybrook Health Sciences Centre 2075 Bayview Avenue Precision Diagnostics and Therapeutics Program Room B204, Toronto, Ontario Canada M4N 3M5 Email: s.

## Abstract

**BACKGROUND:** Many transfusion reactions are not reported to hemovigilance systems, and the features needed to recognize them are often documented only in free-text clinical notes. We sought to develop and evaluate an open-source, multi-agent artificial intelligence (AI) pipeline to extract transfusion reaction features defined by the National Healthcare Safety Network (NHSN) Hemovigilance Module from clinical notes.

**METHODS:** We curated 181 features from the NHSN protocol and developed a multi-stage pipeline that extracted features, their timing, and clinical details from free-text notes. Three open-weight large language models (Qwen 3.6-27B, GPT-OSS-120B, and Kimi K3) were applied to 50 notes from expert-written scenarios, published case reports, and MIMIC-IV discharge summaries. Two physicians independently evaluated every model output in a blinded interface. The primary analysis was restricted to feature occurrences with concordant physician assessments.

**RESULTS:** Physicians agreed on 1,201 of 1,343 co-rated feature occurrences (89.4%; Krippendorff’s *α* 0.73). Precision was above 0.98 for all models. Sensitivity was 0.774 (95% CI 0.745–0.800) for Kimi K3, 0.636 (0.604–0.667) for GPT-OSS-120B, and 0.624 (0.593–0.655) for Qwen 3.6-27B, and specificity ranged from 0.952 to 0.991. Omissions accounted for more than 94% of false negatives. Timing and clinical details were judged supported or reasonable in more than 92% and 97% of assessments, respectively. Results were consistent across note sources and in analyses including discordant assessments.

**CONCLUSION:** Multi-agent AI pipelines provide a feasible method for extracting transfusion reaction signals from clinical notes with high precision. Our work may have applications in active hemovigilance, reaction classification, and big data research.

## 1 INTRODUCTION

All major hemovigilance systems worldwide rely on passive surveillance and reporting of transfusion reactions. [1–3] Studies of active surveillance show that a majority of reactions go undetected. [4–6] Potential reasons include under recognition of signs and symptoms, underreporting from clinical teams to transfusion services, and workload burdens related to processing reaction information. [7, 8] Yet every reaction represents an important signal for blood suppliers to monitor, clinicians to diagnose, patients to understand, transfusion services to track, and researchers to study. Traditionally, a barrier to active detection of reactions has been the diversity and multiplicity of sources that carry facts about transfusion reactions. The increasing use of natural language processing (NLP) and artificial intelligence (AI) technologies to process vast amounts of clinical context present an opportunity for automated detection of reactions. [9, 10]

A key challenge in the detection of transfusion reactions from EHRs is that medical records are voluminous, continuous, complex, and rarely written synoptically or for the purpose of describing transfusion reactions. [9] For instance, a routine nursing or clinical note may describe an unreported transfusion reaction among a myriad of other medical issues and interventions, all of which could potentially contribute to a clinical event. Traditional NLP pipelines have struggled to account for clinical context in coding clinical information, for example, distinguishing affirmative mentions of a clinical feature (“the patient *has hypertension*”) from negated mentions (“the patient *does not have hypertension*”); semantically equivalent expressions (“the patient has a BP of 196/112”); historical or resolved conditions (“the patient *had hypertension, which has since resolved*”); and qualified mentions (“the patient has white-coat hypertension”), as well as other contextual nuances. [11] Foundational large language models (LLMs) can potentially address this limitation by using ‘attention’ mechanisms to encode key facts with awareness of surrounding clinical background, without the need for explicit rules (Table 1). [12–14] As such, LLM pipelines have been successfully applied to extract key findings from radiology evaluations, pathology reports, and procedure notes for the purposes of coding or research. [15–18] Transfusion reactions represent a structurally similar problem, whereby key features may be buried or mixed-in with irrelevant clinical information, requiring specialized context-specific data extraction that can be time-intensive and cost-prohibitive to otherwise perform at scale.

**Table 1.** Glossary of key terms.

| **Term** | **Definition** |
|---|---|
| Natural Language Processing (NLP) | A field of computing that allows software to read and interpret written human language, such as clinical notes |
| Large Language Models (LLMs) | A type of artificial intelligence trained on very large amounts of text that can read a passage and produce written outputs. Here it reads clinical notes and reports the features it finds. |
| AI Agent | AI-based software that can work toward a goal, deciding what steps to take and using tools to carry them out. |
| Pipeline | A sequence of automated processing steps applied to each clinical note to identify and extract relevant clinical features. |
| Feature Extraction | The process of identifying and extracting individual clinical features, such as symptoms or laboratory abnormalities, from clinical notes for subsequent analy-sis. |
| Prompt | The written instructions provided to a language model specifying which clinical features to identify and how to report the extracted information |
| Temperature | A setting that controls how varied a language model's output is for the same input. Lower values make responses more consistent and repeatable, whereas higher values make them more varied. |
| Groq | A commercial platform that provides access to language models over the internet through an API. Here, it was used to run two of the open-weight models: Qwen 3.6-27B and GPT-OSS-120B, which extracted features from the clinical notes. |
| OpenRouter | A commercial platform that provides access to language models over the internet through an API. Here, it was used to run the third open-weight model: Kimi K3. |

In this study we describe the development and transfusion-expert evaluation of an LLM-based pipeline to extract targeted transfusion reaction features from free text medical notes and vignettes. Our aim was to create a generalizable open-source framework and pipeline to extract features relevant for the recognition and classification of transfusion reactions.

## 2 METHODS

### 2.1 Clinical Cases

Clinical notes were sampled from three sources to capture diversity in style, word count, and clinical complexity. The first source was a dataset from a recent study of 36 expert-written transfusion-reaction scenarios reported by Fung et al in the journal Transfusion. [19] The second source was a PubMed Central (PMC) clinical-case dataset (PMC-Patients, version 2) assembled from patient case reports in open-access PMC articles. [20] The third source was MIMIC-IV, a database of de-identified hospital discharge summaries from Beth Israel Deaconess Medical Center in Boston, which represented the most complex and realistic set of clinical notes. [21] For PMC and MIMIC-IV, we used raw-text searches for reaction-related terms to filter notes from each source (1,210 MIMIC-IV notes and 174 PMC cases), followed by large language model (LLM) triage to flag notes likely to describe a genuine transfusion reaction (424 MIMIC-IV notes and 97 PMC cases) and manual review to select cases which described a potential transfusion reaction. These procedures yielded 424 MIMIC-IV notes and 57 PMC cases. From these three sources, a small number of 18 cases (10 Fung et al cases, 5 MIMIC-IV notes, and 3 PMC cases) randomly sampled for development and refinement of the AI extraction pipeline and were not used in the evaluation. From the remaining dataset, we randomly sampled 10 cases from the study by Fung et al., 20 cases from PMC, and 20 cases from the MIMIC-IV case sets (Supplemental §1, §6).

### 2.2 Transfusion Reaction Features

To create a comprehensive list of features relevant for transfusion reaction classification, we used the National Healthcare Safety Network (NHSN) Hemovigilance Module Surveillance Protocol (Version 3.0). The NHSN module provides a detailed, standardized, widely used framework for definition and classification of transfusion reactions. [22, 23] We began by curating a list of all clinical features referenced in the protocol that influence reaction classification, severity, and imputability judgements. These include clinical signs (e.g. hypertension, third heart sound), symptoms (e.g. pruritus, dyspnea), laboratory findings (e.g. elevated lactate dehydrogenase, positive bacterial culture), other diagnostic test results (e.g. radiologic evidence of bilateral infiltrates), and other relevant observations. This yielded a list of 93 distinct clinical features (henceforth called “features”). For features that indicated the presence of a finding, or “pertinent positives”, (e.g. “chest pain”), we also generated a complementary negative feature, or “pertinent negatives” (e.g. “evidence of absence of chest pain”) to detect instances where explicit evidence was available for the absence of a feature. This yielded 88 pertinent-negative features (the remaining 5 features encoded the absence of a finding), giving a final list of 181 features (Supplemental §2, Figure S2.1).

### 2.3 Pipeline Overview

We developed an automated pipeline that takes a free-text clinical note as input and returns a structured list of features as outputs. Each note was processed in three LLM passes for the primary analysis, plus two exploratory stages (Figure 1).

**Figure 1.**
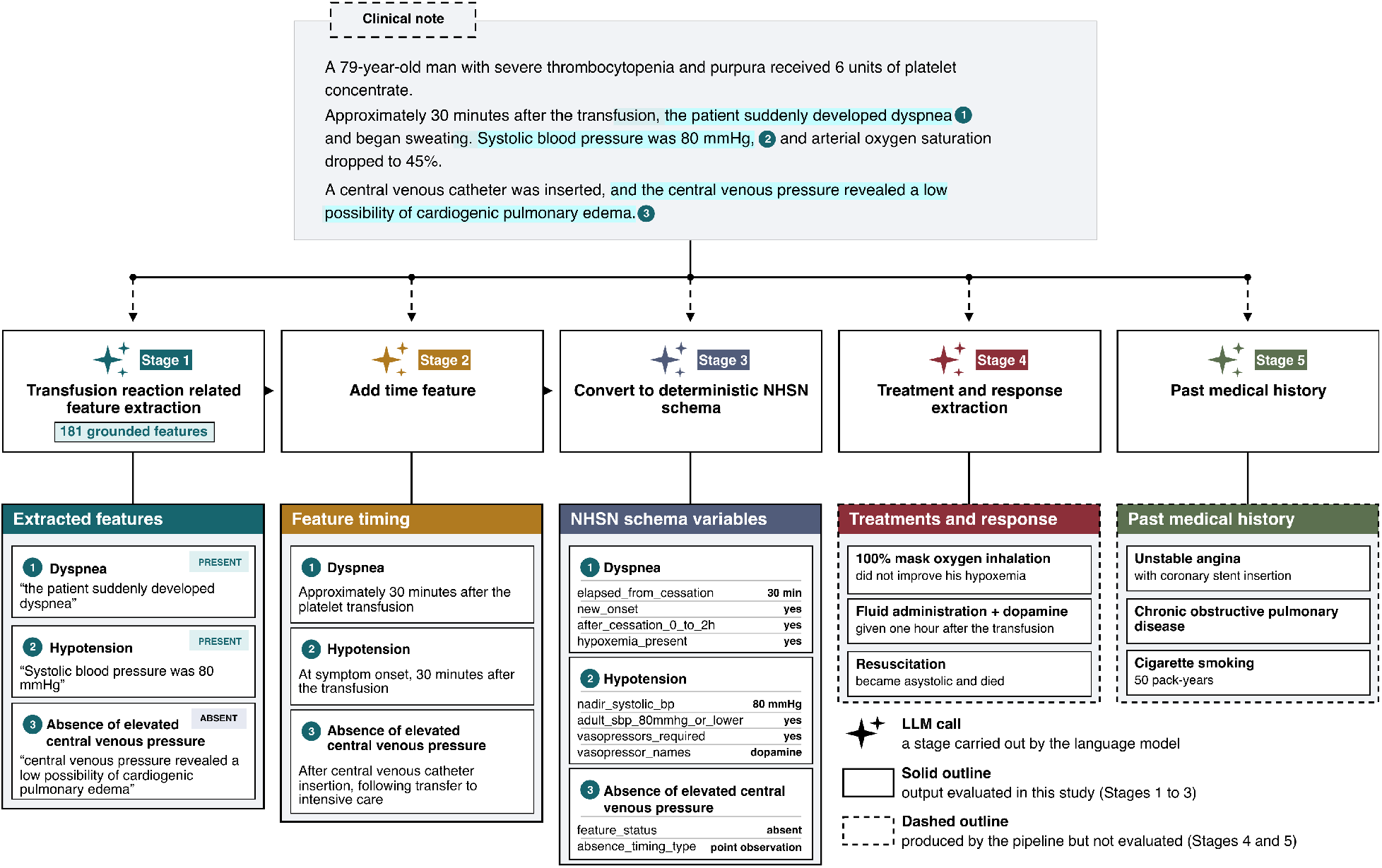
Overview of the large language model pipeline for extracting transfusion reaction features from clinical notes. An example clinical note (top) passes through five stages. Each stage uses calls to the same large language model. Stage 1 extracts clinical features documented in the note. Extraction is restricted to a predefined list of 181 features based on the National Healthcare Safety Network (NHSN) Hemovigilance Module. Each extracted feature is marked present or absent, with a supporting quote from the note. Stage 2 adds the timing of each feature relative to the transfusion. Stage 3 converts features into NHSN schema variables. Stages 4 and 5 extract treatments and past medical history.

The first pass extracted evidence for the presence or absence of clinical features (e.g. “positive blood culture”). The pipeline was designed to restrict LLM outputs to only recognizing findings from the pre-defined list, irrespective of whether they were related to a transfusion event. This restriction was enforced deterministically after Stage 1: each returned feature was matched against the vocabulary, ignoring differences in letter case, hyphenation, brackets and spacing, rewritten to its canonical name, and discarded if it matched no vocabulary item. Pertinent-negative features that repeated an identical evidence phrase were collapsed to a single occurrence. Our purpose in doing so was to separate the task of feature extraction, a fact-finding exercise, from the task of reaction relatedness, which requires more complex medical judgement. For example, if a patient presented with tachypnea on day 1 of admission, and received a transfusion on day 3 of admission, tachypnea was extracted as a feature. The extraction schema and exact LLM prompts are available in Supplemental §4. We additionally extracted pre-defined sub-features for each clinical feature related to timing and pre-specified clinical details. As such, for each feature, the model was instructed to determine whether it was present in the note; if present, the model was instructed to extract (1) the feature’s timing relative to transfusion (e.g. “cough occurred 1 hour after transfusion”), (2) associated clinical details (e.g. “elevated bilirubin 3.1 mg/dL”), and (3) the supporting evidence from the note (e.g. for dyspnea: “patient reported shortness of breath 60 minutes after transfusion”). The primary analysis only examined the presence and absence of features. Where timing or clinical detail was unavailable, the model was instructed to respond with “uncertain”.

Three key properties of feature extraction follow from the above design: first, the extraction target was a closed, controlled vocabulary, restricting the set of possible outputs and limiting unsupported findings; second, the pipeline allowed for extraction to be decomposed into a short chain of single-purpose stages with each performing a narrow task, allowing transparent auditing and troubleshooting of extraction failures. Third, inputs for each stage require correctly structured outputs from the preceding stage, which enforces an internal quality control and reduces noise in the final outputs (Figure 1).

As an exploratory step for future research into reaction classification, and because the information was available in some notes, we additionally extracted past medical history and therapies administered; however, these were not formally evaluated or validated in our study. These extractions are described and sample results presented in Supplemental §4.

### 2.4 Models

All stages involved discrete LLM calls using the same model. The models were Qwen 3.6-27B and GPT-OSS-120B, both served through Groq, and Kimi K3 served through OpenRouter, with full parameters available in the Supplemental. [24–26] Models were chosen on the basis of being open-weight, which is a common requirement in high-risk health data and of representing a range of model sizes. API calls used a temperature of 0 to reduce output variability, except for Kimi K3, which is served at a fixed temperature of 1.0 (Supplemental §3, Table S3.1). All models were run in a structured-output (JSON) mode, so every response conformed to a fixed JSON schema. All models were last accessed on August 20, 2026.

### 2.5 Evaluation Interface

We developed a custom platform to support blinded, asynchronous physician evaluation of clinical feature extractions from the three LLMs (Figure 2). Evaluators received a tutorial on the task and accessed a web-based interface using unique credentials. Within the interface, each evaluator completed a test task to familiarize with the interface, and completion granted access to a list of evaluation tasks. Each evaluation task consisted of model-identified features organized into three randomized, blinded model tabs.

**Figure 2.**
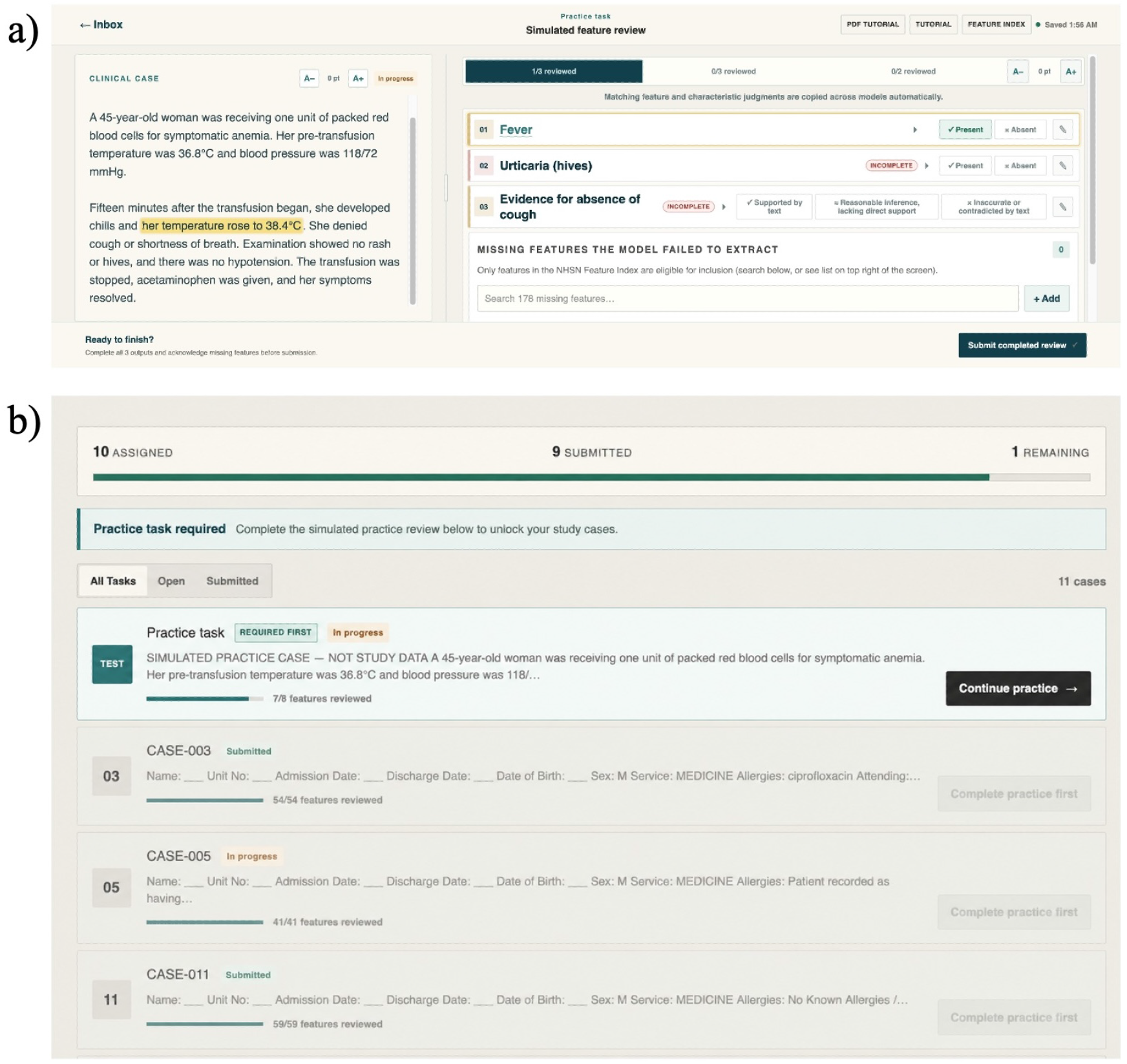
Evaluation interface depicting a) case evaluation interface and b) case task list

Evaluators classified features as present or absent and assessed timing and clinical features marked present. For each model tab, evaluators also had the opportunity to select features present in the note and missing in the extraction, termed “missing features”.

To improve consistency, applicable judgments about presence/absence of features were propagated across model tabs when the same feature occurred in multiple model outputs, which the evaluators had the ability to modify. Evaluations occurred over 6 weeks and data were exported for analysis as comma-separated files. Additional details are available in Supplemental §5.

### 2.6 Data Analysis

We evaluated all three models (Qwen 3.6-27B, GPT-OSS-120B, and Kimi K3) on the same 50 clinical notes, which comprised 10 Fung et al cases, 20 PMC case reports, and 20 MIMIC-IV notes, producing 150 model-note evaluations in total. Two reviewers independently assessed every model’s output, judging for each feature the model reported whether it was accurately identified based on the clinical note, and recording any clinically relevant feature the model had omitted. Descriptive analyses reported mean and standard deviation for normally distributed and median and interquartile range for skewed variables. The primary analysis was restricted to features on which both reviewers agreed; features with discordant assessments were excluded from the primary analysis and evaluated separately in a supplementary analysis (Supplemental §7, Table S7.1). Each reviewer’s judgements were counted separately, and omissions recorded by either reviewer were retained.

We used confusion matrices to summarize how well each model distinguished present from absent features. Features correctly identified as present or absent were counted as true positives or true negatives, with true negatives being correctly identified pertinent negatives; features incorrectly identified as present were counted as false positives; and false negatives were features present in the note that the model either marked as absent (negated misses) or did not report at all (omissions). From these counts we calculated precision, sensitivity, specificity, and F1 for each model, both overall and by note source, with 95 percent confidence intervals for sensitivity and specificity using the Wilson score method. For features the reviewers confirmed as present, we also assessed how accurately each model captured timing and associated clinical details. Each NHSN schema variable extracted in Stage 3 was rated separately by each reviewer (blank ratings were excluded), with the reviewer rating the extracted information (e.g. “no S3 heard”) as supported by the note (“Normal S1, S2, no S3”), a reasonable inference based on clinical context (“Normal heart sounds present”), or inaccurate (“S3 present”); from these ratings we report a strict rate (supported only) and a lenient rate (supported or reasonable inference). To measure agreement beyond chance, we calculated Gwet’s AC1 with 95 percent confidence intervals alongside the observed percentage agreement. Finally, we described how many features each model reported per note as the median and interquartile range, overall and by note source. We also counted false positives and false negatives in each physician evaluation of each case, by model (Figure 3). This study is reported in accordance with the TRIPOD-LLM reporting guideline. [27] All analyses were performed in Python (version 3.13.3).

**Figure 3.**
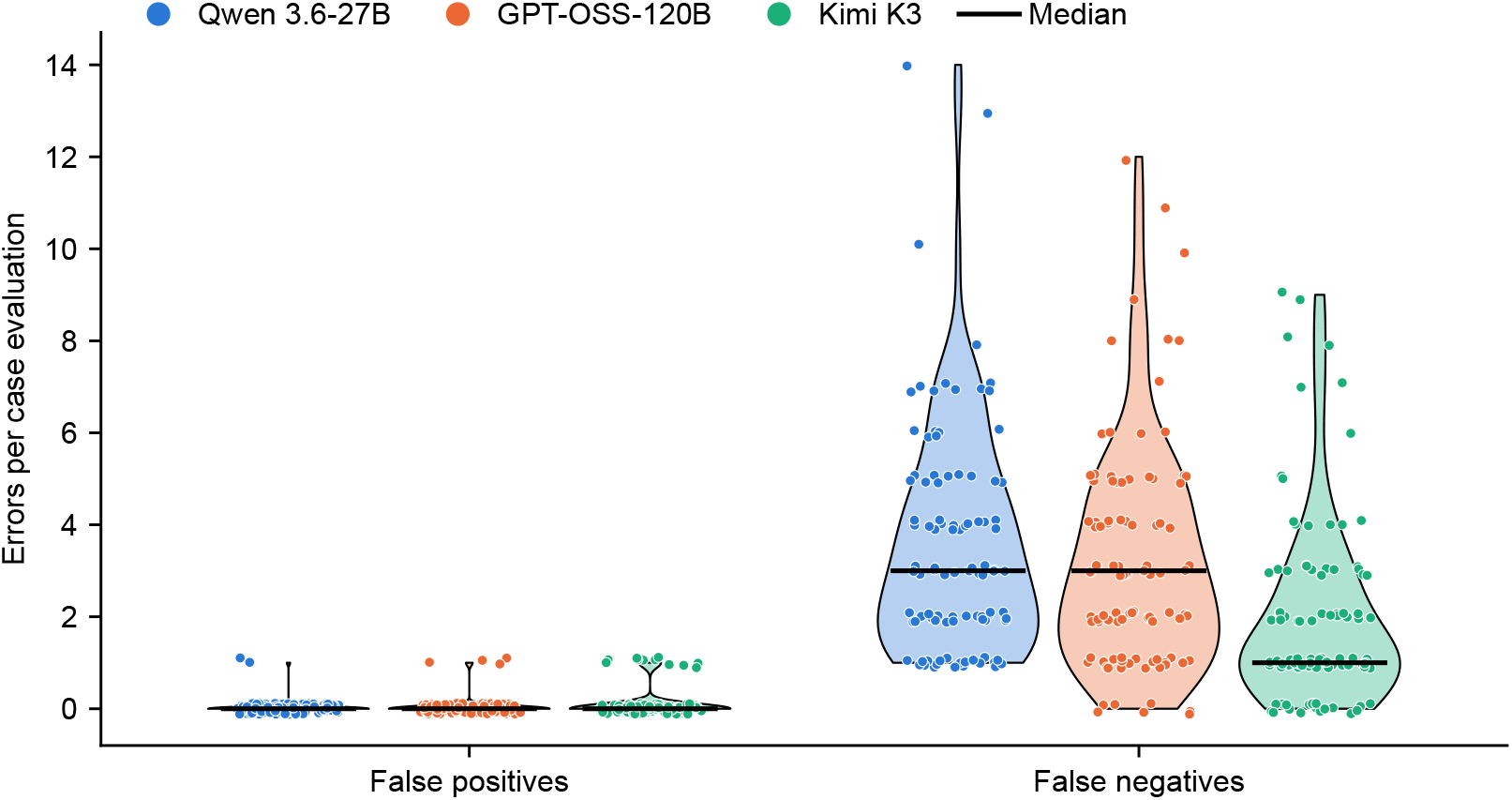
False positives and false negatives per case evaluation, by model. Each dot represents one physician’s evaluation of one case (50 cases, two reviewers each; 100 evaluations per model). Violins show the distribution of error counts; black lines mark the median. Only feature occurrences with concordant physician assessments are included. False negatives combine omissions and features incorrectly marked absent.

## 3 RESULTS

We evaluated three large language model pipelines (Qwen 3.6-27B, GPT-OSS-120B, and Kimi K3) using 50 clinical notes from three sources: Fung et al, PMC case reports, and MIMIC-IV notes. Every extracted feature was compared against a physician reference standard, in which two reviewers independently judged whether each feature was present, absent, or not reported. The two physician reviewers co-rated 1,343 feature occurrences and agreed on presence or absence for 1,201 (89.4%; Krippendorff’s *α* 0.73, 95% CI 0.69–0.77). The 142 (10.6%) discordant occurrences were excluded from the primary analysis (Supplemental §7, Table S7.1d). When the analysis was repeated with all 1,343 co-rated occurrences, including the discordant ones, the ranking of the three models was unchanged (Supplemental §7, Table S7.1).

### 3.1 Overall Pipeline Performance

All three models achieved high precision (above 0.98), indicating that features identified as present were generally confirmed by physician reviewers (Table 2). The principal difference in performance was observed in sensitivity, with Kimi K3 identifying a greater proportion of documented clinical features (0.77, 95% CI 0.75–0.80) than GPT-OSS-120B (0.64, 95% CI 0.60–0.67) and Qwen 3.6-27B (0.62, 95% CI 0.59–0.66).

**Table 2.** Model performance against the physician reference standard (concordant feature occurrences). Sensitivity and specificity with Wilson 95% confidence intervals.

| **Model** | **Precision** | **Sensitivity (95% CI)** | **Specificity (95% CI)** | **F1** |
|---|---|---|---|---|
| Qwen 3.6-27B | 0.99 | 0.62 (0.59–0.66) | 0.99 (0.97–0.99) | 0.77 |
| GPT-OSS-120B | 0.99 | 0.64 (0.60–0.67) | 0.96 (0.91–0.99) | 0.78 |
| Kimi K3 | 0.98 | 0.77 (0.75–0.80) | 0.95 (0.92–0.97) | 0.87 |

Sensitivity estimates for GPT-OSS-120B and Qwen 3.6-27B were similar, with overlapping confidence intervals. Specificity was 0.99 (95% CI 0.97–0.99) for Qwen 3.6-27B, 0.96 (0.91–0.99) for GPT-OSS-120B, and 0.95 (0.92–0.97) for Kimi K3. Kimi K3 also achieved the highest F1 score (0.87), compared with GPT-OSS-120B (0.78) and Qwen 3.6-27B (0.77). When discordant occurrences were included, the ranking of models was unchanged (sensitivity 0.78, 0.65, and 0.62 for Kimi K3, GPT-OSS-120B, and Qwen 3.6-27B, respectively; Supplemental §7, Table S7.1b).

### 3.2 Model Performance across clinical note sources

To examine whether the pipelines performed consistently across different forms of clinical documentation, we evaluated feature identification separately for Fung et al cases, PMC case reports, and MIMIC-IV notes (Table 3). Sensitivity varied across clinical note sources, although the relative performance of the three models remained consistent (Table 3). Kimi K3 achieved the highest sensitivity across all sources, ranging from 0.68 in Fung cases to 0.82 in PMC case reports. GPT-OSS-120B and Qwen 3.6-27B showed comparable, but consistently lower, sensitivity across sources. All three models demonstrated their lowest sensitivity in Fung cases, suggesting greater difficulty in recovering documented clinical features from these cases. Specificity remained high across clinical note sources (0.82–1.00), with several models achieving perfect specificity in subsets containing relatively few physician-confirmed absent features. Pooled across models, the median number of features correctly identified per note was 3 (IQR 2–4.75) for Fung et al cases, 6 (4–8) for PMC reports, and 7.5 (3–11) for MIMIC-IV notes (Supplemental §7, Table S7.3).

**Table 3.** Model performance across the three case sources.

| **Case source (n notes)** | **Model** | **Sensitivity** | **Specificity** |
|---|---|---|---|
| Fung et al Cases (10) | Qwen 3.6-27B | 0.54 | 0.86 |
|  | GPT-OSS-120B | 0.58 | 1.00* |
|  | Kimi K3 | 0.68 | 0.82 |
| PMC reports (20) | Qwen 3.6-27B | 0.65 | 1.00* |
|  | GPT-OSS-120B | 0.67 | 1.00* |
|  | Kimi K3 | 0.82 | 0.95 |
| MIMIC-IV notes (20) | Qwen 3.6-27B | 0.62 | 1.00* |
|  | GPT-OSS-120B | 0.63 | 0.95 |
|  | Kimi K3 | 0.76 | 0.97 |

*\*A specificity of 1.00 means no features were wrongly reported as present. This value derives from only a small number of features that the physicians marked absent, and should be interpreted as “no errors of commission seen in this case-model configuration” rather than as precise estimates of true specificity.*

### 3.3 Composition of errors

To further characterize the differences in feature identification, we examined the types of disagreement between model outputs and physician assessments. Across all three models, most false negatives resulted from omissions, where clinically relevant features were not reported, rather than negated misses, where features were incorrectly marked as absent (Table 4). Omissions accounted for 94.9% of false negatives in Qwen 3.6-27B, 100% in GPT-OSS-120B, and 99.0% in Kimi K3. Kimi K3 had the fewest false negatives overall (n=196), compared with GPT-OSS-120B (n=317) and Qwen 3.6-27B (n=354). False positives were uncommon across all three models, with 2 for Qwen 3.6-27B, 4 for GPT-OSS-120B, and 12 for Kimi K3. Per case evaluation, the median number of false negatives was 3 (IQR 2–5) for Qwen 3.6-27B, 3 (1–4.25) for GPT-OSS-120B, and 1 (1–3) for Kimi K3, while false positives had a median of 0 and a maximum of 1 for all models (Figure 3). Among features reported as absent, physicians confirmed absence in 108 of 117 (92.3%) for Qwen 3.6-27B, 50 of 50 (100%) for GPT-OSS-120B, and 118 of 119 (99.2%) for Kimi K3. Frequently missed features included laboratory and definitional criteria such as “unexplained clinical illness consistent with infection” (missed in 10 of 11, 8 of 10, and 3 of 9 occurrences, respectively) (Supplemental §7, Tables S7.2a–d).

**Table 4.**
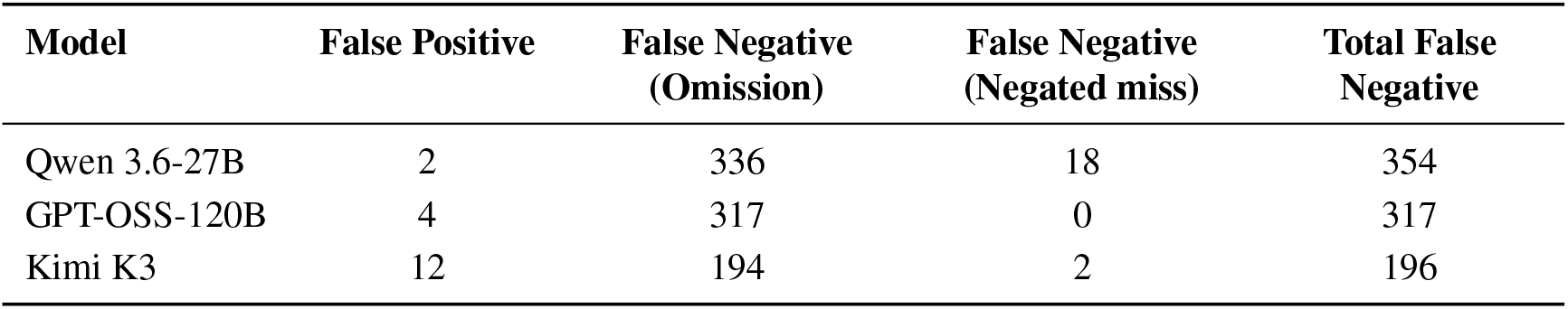
Breakdown of errors by type for each model. Negated misses = the feature was recorded as absent; Omissions = the feature was not mentioned at all. Total false negatives = negated misses + omissions.

| **Model** | **False Positive** | **False Negative (Omission)** | **False Negative (Negated miss)** | **Total False Negative** |
|---|---|---|---|---|
| Qwen 3.6-27B | 2 | 336 | 18 | 354 |
| GPT-OSS-120B | 4 | 317 | 0 | 317 |
| Kimi K3 | 12 | 194 | 2 | 196 |

### 3.4 Quality of Extracted Information

Beyond identifying clinical features, we evaluated the accuracy of their associated timing and descriptive details. Descriptive details were extracted more accurately than timing information across all three models (Table 5). Under the strict definition, detail accuracy was highest for GPT-OSS-120B (93.5%), compared with Kimi K3 (92.4%) and Qwen 3.6-27B (91.2%). When reasonable inferences were included, Kimi K3 achieved the highest accuracy (98.2%), with slightly lower values for Qwen 3.6-27B (97.9%) and GPT-OSS-120B (97.4%). Timing accuracy was lower across models, with Kimi K3 achieving the highest rates under both strict (88.7%) and lenient (97.6%) definitions, compared with GPT-OSS-120B (88.4% and 95.3%) and Qwen 3.6-27B (81.8% and 92.7%). Inaccurate assessments accounted for at most 7.3% of timing evaluations and 2.6% of detail evaluations.

**Table 5.** Accuracy of extracted timing and descriptive detail. Supported = judged exactly accurate; Reasonable inf. = a clinically reasonable inference; Inaccurate = incorrect. Strict % counts only Supported entries; Lenient % also credits Reasonable inferences.

| **Model** | **Assessment** | **Supported** | **Reasonable inf.** | **Inaccurate** | **Strict %** | **Lenient %** |
|---|---|---|---|---|---|---|
| Qwen 3.6-27B | Timing | 1036 | 138 | 92 | 81.8% | 92.7% |
|  | Details | 2152 | 159 | 49 | 91.2% | 97.9% |
| GPT-OSS-120B | Timing | 790 | 62 | 42 | 88.4% | 95.3% |
|  | Details | 1827 | 75 | 51 | 93.5% | 97.4% |
| Kimi K3 | Timing | 1415 | 143 | 38 | 88.7% | 97.6% |
|  | Details | 2556 | 160 | 51 | 92.4% | 98.2% |

### 3.5 Model-Physician Agreement

To assess overall agreement between model outputs and physician assessments, we calculated Gwet’s AC1, accounting for agreement expected by chance. Kimi K3 showed the highest chance-corrected agreement with physician reviewers (Gwet’s AC1 0.68, 95% CI 0.63–0.72), with an observed agreement of 81.3%. Agreement was lower for GPT-OSS-120B (AC1 0.46, 95% CI 0.40–0.52) and Qwen 3.6-27B (AC1 0.44, 95% CI 0.39–0.50), with observed agreement of 67.1% and 69.3%, respectively. The confidence intervals for GPT-OSS-120B and Qwen 3.6-27B overlapped, whereas neither overlapped with that of Kimi K3.

## 4 DISCUSSION

We developed and evaluated a multi-stage AI pipeline to extract standardized transfusion reaction features from three sources of free-text clinical notes and compared performance across three open-weight LLMs. Models achieved modest sensitivity, detecting about three-quarters of all features present, and very high specificity. The largest model (Kimi K3) generally yielded the best performance, and all models achieved higher sensitivity on published case reports (PMC) and real-world notes (MIMIC-IV) than on expert-written scenarios. Sensitivity was modest, with models discovering 2/3 to 3/4 of features, and models rarely reported features that were not present (specificity *≥*0.95). Models were also able to extract additional details about the timing and clinical descriptions of reported features, with greater than 90% accuracy across all model/note combinations. Our results demonstrate the technical feasibility of clinical note extraction on a task that is typically time intensive and requires clinical expertise.

Several findings deserve nuanced interpretation. The modest sensitivity in our study partly reflects inherent complexity and potential overlap of reactions characteristics described in the NHSN. For instance, the pairs ‘hematuria’ and ‘hemoglobinuria’, and, ‘acute or worsening respiratory distress’ and ‘respiratory distress’, appear among the top 10 most commonly missed features, and some models extracted one feature but not the other, while human experts tended to code both as accurate or reasonable extractions (Supplemental §7). Prior work has also reported that LLMs may generate unsupported content when information is absent from the source document. [28] The high precision in our study suggests that restricting outputs to a closed vocabulary and requiring verbatim supporting evidence limited such errors. We found that Kimi K3 performed best, followed by GPT-OSS-120B and Qwen 3.6-27B, which is expected and follows the order of parameter size of these models. Contrary to our expectations, we found that models performed better on the most complex, but also most realistic, clinical notes (MIMIC-IV) than on the expert-written scenarios, potentially indicating that rich clinical context improves LLM performance.

Our work extends previous research, where multi-agent LLM pipelines have been applied to extract structured information from free-text clinical documentation, including radiology reports, pathology reports, and hospital discharge summaries. [13] GPT-4 extracted lesion information from lung cancer CT reports with 98.6% accuracy, [29] GPT-3.5 classified lung cancer pathology reports with 89% accuracy, [28] and a locally deployed open-weight model identified liver cirrhosis in MIMIC-IV medical histories with 100% sensitivity and 96% specificity. [30] Within transfusion medicine, automated surveillance has largely relied on rule-based electronic health record algorithms targeting individual reactions such as transfusion-related acute lung injury and transfusion-associated circulatory overload, [31] and recent work has evaluated LLMs on standardized transfusion reaction scenarios. [19] To our knowledge, our study is among the first to evaluate multi-agent LLM extraction of a standardized transfusion reaction feature list from real-world clinical notes.

Our findings provide useful insights for modern hemovigilance initiatives. Passive reporting systems are time intensive and often fail to capture most transfusion reactions. In one study of electronic surveillance, only 11 of 78 algorithm-detected cases of transfusion-related acute lung injury and 5 of 45 cases of transfusion-associated circulatory overload had been reported to the blood bank. [31] Structured, evidence-linked features extracted from clinical notes could be mapped to NHSN case definitions, severity, and imputability criteria, allowing transfusion services to identify potential reactions without relying on clinician recognition and reporting. Because each extracted feature is linked to verbatim evidence from clinical notes, outputs can be audited by transfusion medicine physicians or transfusion safety officers, expanding capacity while providing expert oversight.

### Strengths

Our study has several strengths. Clinical notes were drawn from three sources that differed in style, length, and complexity, including expert-written scenarios, published case reports, and real-world hospital discharge summaries. All three models were open-weight, which ensures feasibility of local deployment in health data environments where sensitive data cannot be shared with external services. Every model output was independently assessed by two physicians using a blinded, randomized evaluation interface, with acceptable inter-rater agreement (89.4% observed agreement; Krippendorff’s *α* 0.73), supporting the reliability of the reference standard. The feature vocabulary and extraction stages were pre-specified from the NHSN protocol before evaluation to ensure extractions were constrained and clinically useful. The pipeline decomposed extraction into single-purpose stages with evidence-linked outputs, allowing errors to be traced to a specific stage and feature, and all prompts and code are available for replication (Supplemental §4, §8). Automated extraction can reduce the manual chart review required to identify reaction features and is readily scalable to large clinical datasets.

### Limitations

The pipeline assumed a single transfusion reaction per note and did not separate multiple transfusions, multiple reactions, or symptoms that recurred at different time points. Ambiguous or overlapping clinical descriptions were a source of error; for example, rib pain was coded as back pain, and hematuria and hemoglobinuria were among the most frequently missed features and can be refined in future work. Timing was more difficult to extract than clinical details, with strict timing accuracy of 81.8% to 88.7% across models. This is partly because we used only data from clinical notes, and retrieving laboratory and transfusion data directly from structured tables would yield more complete and accurate data on timing and transfusions. The models performed worst on synthetic clinical summaries, and the pipeline will require additional tuning prior to use with existing hemovigilance databases that contain only reaction summary data. This also applies to real-world notes that have not been pre-processed for research as MIMIC-IV notes have, and may carry duplicated content, formatting noise, and boilerplate text that may lack informational value and must be pre-processed prior to content extraction and use in AI pipelines. Physician validation of feature extractions was demanding, taking about 10-20 minutes per task, which limited the sample size of evaluations. Sensitivity was calculated from features physicians recorded as missing, and specificity was calculated among features the models reported; neither represents a classic diagnostic accuracy estimate. Each model was run once per note and on the lowest available temperature setting, however LLM responses show stochasticity. Larger models such as Kimi K3, while more accurate, also incur higher computational and environmental costs, which we did not measure.

### Implications

Several potential extensions arise from our work. First, extracted features can serve as inputs to semi-deterministic pipelines for transfusion reaction classification, in which LLMs perform fact-finding and NHSN criteria are applied by rules. Second, the pipeline can support active hemovigilance by flagging notes with potential reactions for review by transfusion services and blood suppliers. Third, automated feature extraction can be used to code transfusion reaction features in large clinical datasets for research and as inputs to machine learning models. Fourth, the approach can be adapted for adverse event monitoring in clinical trials of blood products. Finally, future research should validate the extraction of treatments and past medical history, verify extracted findings against structured laboratory and vital sign data, and add reaction classification as the next stage of the pipeline.

## 5 CONCLUSION

We developed and evaluated an LLM pipeline that extracts NHSN-defined transfusion reaction features from free-text clinical notes and found that open-weight models identified features with high precision, with omission of documented features as the main limitation. Our open framework provides a foundation for automated, auditable detection and classification of transfusion reactions to support active hemovigilance.

## Data Availability

MIMIC-IV-Note discharge summaries are available to credentialed users via PhysioNet (https://doi.org/10.13026/1n74-ne17) and cannot be redistributed by the authors. PMC-Patients case reports and the Fung et al. scenarios are publicly available from their original publications. All pipeline code, prompts, the feature vocabulary, and the identifiers of the notes used in this study are available at https://github.com/Ayu-98-kumari/Multi-Agent-Generative-Artificial-Intelligence-for-Transfusion-Reaction-Feature-Extraction.

https://doi.org/10.13026/1n74-ne17

## ABBREVIATIONS

AC1: agreement coefficient 1
AI: artificial intelligence
API: application programming interface
CSV: comma-separated values
EHR: electronic health record
LLM: large language model
MIMIC-IV: Medical Information Mart for Intensive Care IV
NHSN: National Healthcare Safety Network
NLP: natural language processing
PMC: PubMed Central

## ACKNOWLEDGMENTS

None.

## CONTRIBUTIONS

AK, SR, and AA completed the first draft. AK and AA performed data analysis. All authors reviewed the manuscript draft.

## ETHICS STATEMENT

All data were de-identified. MIMIC-IV discharge summaries were accessed under the PhysioNet Credentialed Health Data Use Agreement; collection and de-identification of MIMIC-IV were approved by the Institutional Review Boards of Beth Israel Deaconess Medical Center and the Massachusetts Institute of Technology, with a waiver of informed consent. MIMIC-IV notes were processed in accordance with PhysioNet guidance on the responsible use of credentialed data with online services. Notes were sent to the large language models through application programming interfaces (Groq and OpenRouter) configured for zero data retention, with no use of submitted data for model training and no human review. PMC-Patients case reports and the Fung et al. scenarios are publicly available. As the study used only de-identified data, it did not require ethics review.

## DATA AVAILABILITY

MIMIC-IV-Note discharge summaries are available to credentialed users via PhysioNet (https://doi.org/10.13026/1n74-ne17) and cannot be redistributed by the authors. PMC-Patients case reports and the Fung et al. scenarios are publicly available from their original publications. [19, 20] All pipeline code, prompts, the feature vocabulary, and the identifiers of the notes used in this study are available in the code repository (Supplemental §8).

## Appendix

Multi-Agent Generative Artificial Intelligence for Transfusion Reaction Feature Extraction

### §1 Case-Selection Pipeline

Clinical notes were drawn from three sources.

1. MIMIC-IV hospital discharge summaries [21]
2. PMC-Patients published case reports [20]
3. Publication on AI for transfusion reactions by Fung et al. [19] (public domain)

For sources #1 and #2, we began by using a deterministic word search for the term ‘transfusion-reaction-terms’ and their acronyms (Table S6.1) to filter notes. This was followed by an LLM-based flagging for cases likely to contain a transfusion reaction. In a final step, the senior author SR manually filtered through all cases to select those with a potential transfusion reaction. The procedure is described in detail in §6 below.

All cases in source #3 were synthetically generated by the authors of that study for transfusion reaction evaluation and were thus potential transfusion reactions by design.

Additional details are provided below:

1. **MIMIC-IV discharge summaries.** The de-identified MIMIC-IV note table (discharge.csv.gz) contains 331,793 discharge summaries, with the narrative read from the note’s text field. Because the table is distributed as a single gzip-compressed CSV, it was streamed in fixed 5,000-row chunks and never decompressed to disk or held fully in memory, keeping the peak memory footprint constant regardless of file size. MIMIC-IV is used under the PhysioNet credentialed-access Data Use Agreement (DUA) and cannot be redistributed.
2. **PMC-Patients case reports.** PMC-Patients is an open-access collection of 250,294 patient case reports extracted from published PubMed Central articles, distributed as a single JSON array of approximately 837 MB. Before screening, the array was converted to a flat CSV by streaming it one record at a time, so that peak memory remained flat irrespective of the file’s size. Every column was preserved; nested fields such as age (stored in the source as a list of value/unit pairs) were serialized to JSON strings so that no structure was lost and the original value could be recovered by parsing the cell. The narrative was read from the patient field. PMC-Patients is open access and carries no DUA.
3. **Clinical scenarios from Fung et al study.** The case set comprised 36 hypothetical posttransfusion adverse-event scenarios originally developed for a 2014 validation study of the CDC National Health-care Safety Network (NHSN) Hemovigilance Module definitions. The scenarios were constructed to test application of standardized transfusion-reaction criteria and were independently classified by a transfusion medicine expert panel, whose determinations served as the reference standard. All but one scenario had a single intended classification; case 25 contained two simultaneous reactions, yielding 37 possible transfusion reactions across the 36 cases. Six scenarios intentionally did not satisfy the criteria for any NHSN adverse-event category and were therefore intended for classification as no reaction.

### §2 Controlled Feature Vocabulary Creation

Case definitions for following reactions were extracted from CDC/NHSN Protocol Version 3.0

| **No.** | **Reaction (as used to build the vocabulary)** |
|---|---|
| 1 | Transfusion-transmitted infection (TTI) |
| 2 | Acute hemolytic transfusion reaction (AHTR) |
| 3 | Transfusion-associated circulatory overload (TACO) |
| 4 | Transfusion-related acute lung injury (TRALI) |
| 5 | Allergic transfusion reaction |
| 6 | Delayed hemolytic transfusion reaction (DHTR) |
| 7 | Delayed serologic transfusion reaction (DSTR) |
| 8 | Febrile non-hemolytic transfusion reaction (FNHTR) |
| 9 | Hypotensive transfusion reaction (HTR) |
| 10 | Post-transfusion purpura (PTP) |
| 11 | Transfusion-associated dyspnea (TAD) |
| 12 | Transfusion-associated graft-versus-host disease (TAGVHD) |

A list of clinical features (symptoms, signs, clinical investigations, and syndromes) was extracted from case definitions. The features were then pooled and the list was deduplicated (e.g. ‘shortness of breath’ and ‘dyspnea’ were collapsed into dyspnea). A negative counterpart was then created by appending the phrase ‘Evidence for absence of <feature> ‘, for example, ‘Evidence for absence of fever’. This was done to allow us to explicitly instruct the model to detect pertinent negatives when mentioned, in addition to pertinent positives, during the extraction steps. Because some primary features extracted from the NHSN were already phrased in the negative, redundant double negatives were removed.

**Figure S2.1.**
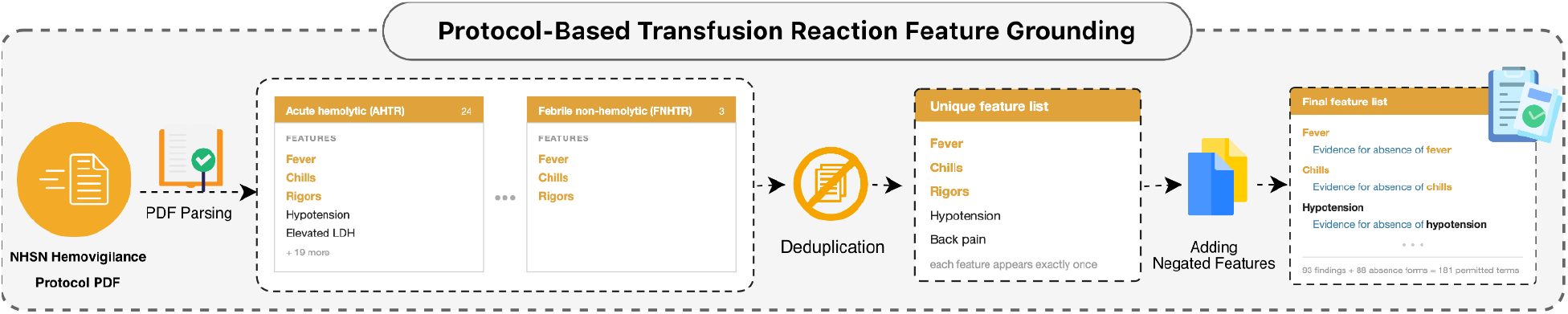
Construction of the controlled feature vocabulary. Clinical features were extracted from the case definitions of 12 transfusion reactions in the NHSN Hemovigilance Module protocol, pooled, and deduplicated, giving 93 features. A pertinent-negative form (“Evidence for absence of…”) was added for 88 features, giving 181 features in total.

### §3 Models and Parameters

All three models were accessed through programmatic, stateless API calls rather than interactive chat sessions. The same model was used for every stage of a given pipeline run. All calls used JSON output mode, so each response conformed to a fixed JSON schema. Responses truncated by the output-token limit were rejected rather than repaired. Transient and rate-limit errors were retried up to 10 times with exponential backoff (initial wait 1 s, factor 2, maximum 60 s). Outputs were cached per note so that each stage could be audited and re-run reproducibly.

**Table S3.1.** Model and inference parameters.

| **Model** | **Provider** | **Model identifier** | **Temperature** | **Reasoning effort** | **Maximum output tokens** |
|---|---|---|---|---|---|
| Qwen 3.6-27B | Groq | qwen/qwen3.6-27b | 0 | Disabled* | 16,384 |
| GPT-OSS-120B | Groq | openai/gpt-oss-120b | 0 | Medium | 50,000 |
| Kimi K3 | OpenRouter (Moonshot AI; provider fallback disabled) | moonshotai/kimi-k3 | 1.0† | Low | 20,000 |

\*Reasoning was disabled for Qwen 3.6-27B because its reasoning output is incompatible with JSON output mode. †Kimi K3 is served at a fixed temperature of 1.0; this was the minimum available setting.

### §4 Extraction Stages and Prompts

Each clinical note in the study cohort was processed in 3 extraction stages for the primary analysis, and 2 additional stages for exploratory work. All model outputs were structured JSONs. The model was instructed to quote direct evidence from each clinical note for each extraction.

- **Stage 1, Extracting NHSN features.** Given the note and the 181-feature vocabulary, the model returned each supported feature with its evidence span. The feature set was frozen after stage 1, and used in subsequent stages.
- **Stage 2, extracting timing information for each feature.** Given the note and the features extracted in stage 1, the model extracted temporal details. The model was instructed to express time in relation to the nearest transfusion when available, for example, “6.25 hours after completion of the apheresis platelet transfusion”. Features with no attributable time are marked *uncertain*.
- **Stage 3, extracting additional details for each feature.** For each feature extracted in stage 1, the model was given the clinical note, the feature with its evidence, and that feature’s menu of NHSN Hemovigilance Module variables, with the definition, response type, allowed values, and units of each variable. The model was called separately for timing variables and for details variables, and returned only the variables supported by the note; for example, “vomiting 90 minutes after the transfusion ended” was returned as an elapsed time from transfusion cessation of 90 minutes. Only variables in the feature’s menu were retained. For pertinent negatives, only timing variables were filled by the model, and the details variable was taken from the stage 1 evidence.
- **Stage 4, extracting treatments.** The model extracted the treatments administered and, where stated, their response, timing relative to transfusion, and whether each treats a feature on the list.
- **Stage 5, extracting past medical history.** An independent stage that was given only the clinical note and listed pre-existing conditions, chronic illnesses, and allergies as a list.

| Stage | Input | Output (JSON schema) |
|---|---|---|
| 1: Features + evidence | Note + 181-feature vocabulary | List of {feature, evidence}; absences as “Evidence for absence of …”. |
| 2: Time | Note + numbered Stage-1 features | {“times”: [{“index”, “time”}]}, one per feature; “uncertain” if none. |
| 3: NHSN schema variables (timing and details) | Note + one positive feature (name + evidence) + that feature’s Timing/Details variable menu | {“variables”: [{“variable_id”, “value”, “unit”}]} per feature per group, merged onto the feature as a nested schema object. |
| 4: Treatments | Note + all positive features (name + evidence + time) | {“treatments”: [{“treatment”, “response”, “timing”, “feature_list”}]}. |
| 5: Past medical history | Note only | {“past_medical_history”: [{“condition”, “detail”}]}. |

Limitations:

- Multiple transfusions are not easily separable in the current pipeline
- Multiple transfusion reactions are not easily disaggregated
- Multiple start/stops within a single transfusion are not easily detected or separately flagged

#### Stage 1: Features and Evidence

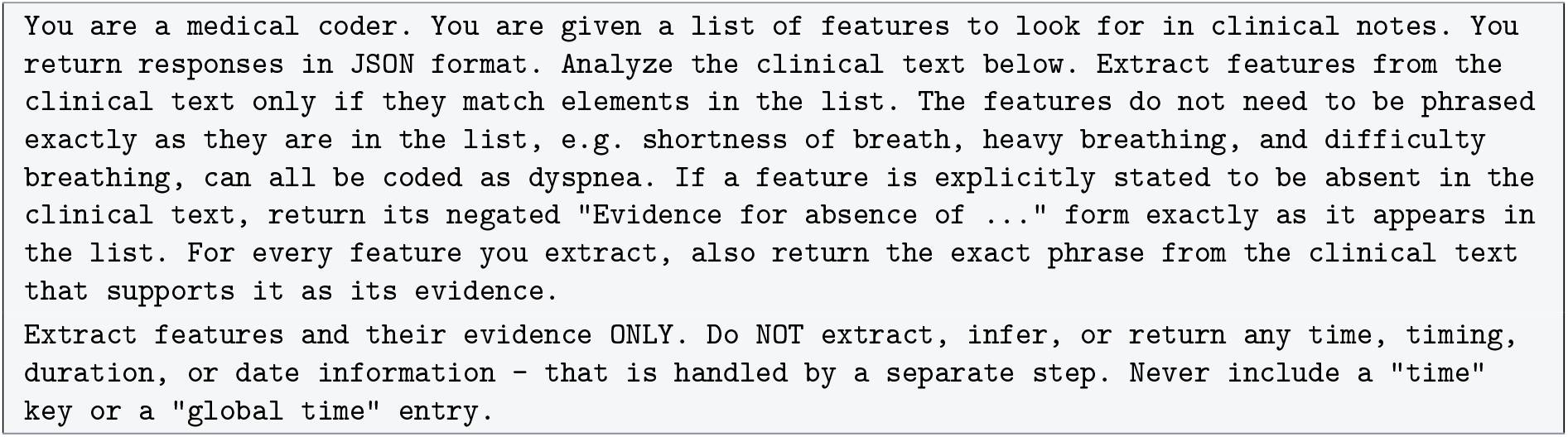

#### Stage 2: Time

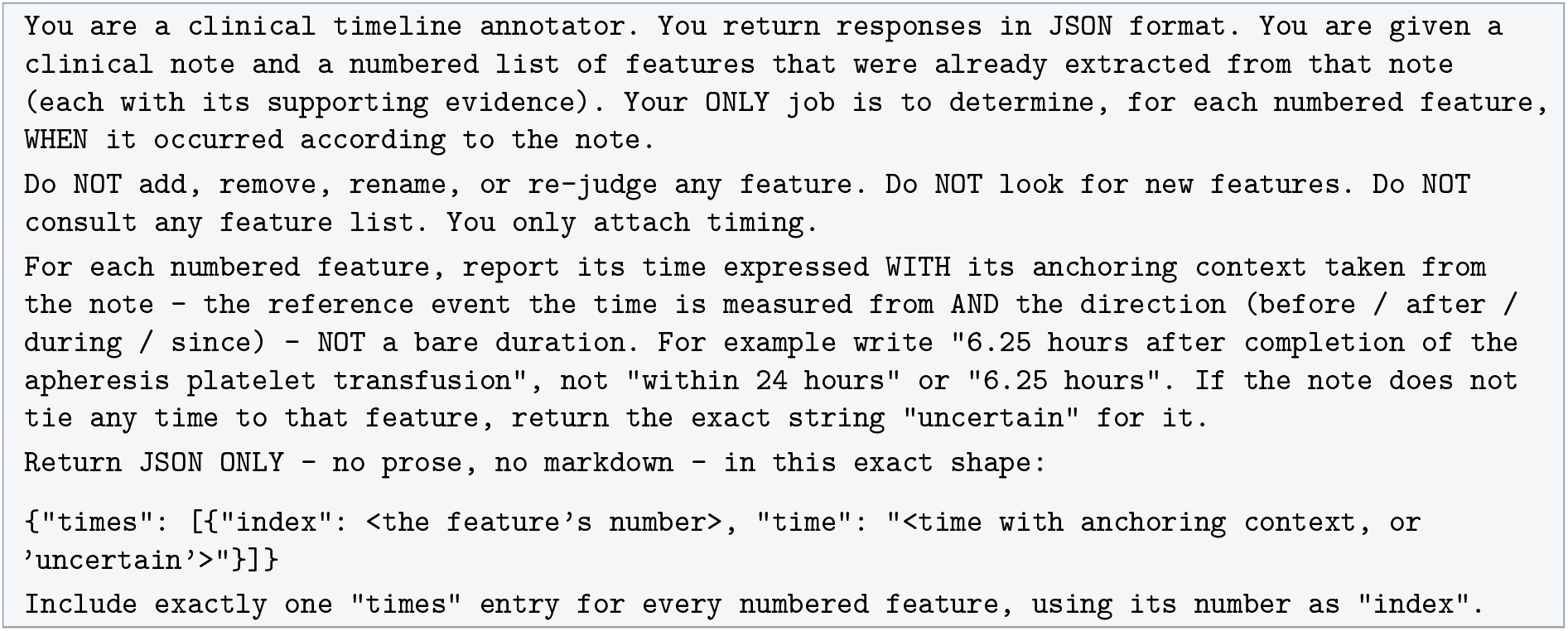

#### Stage 3: NHSN Schema Fill-in (Timing Group)

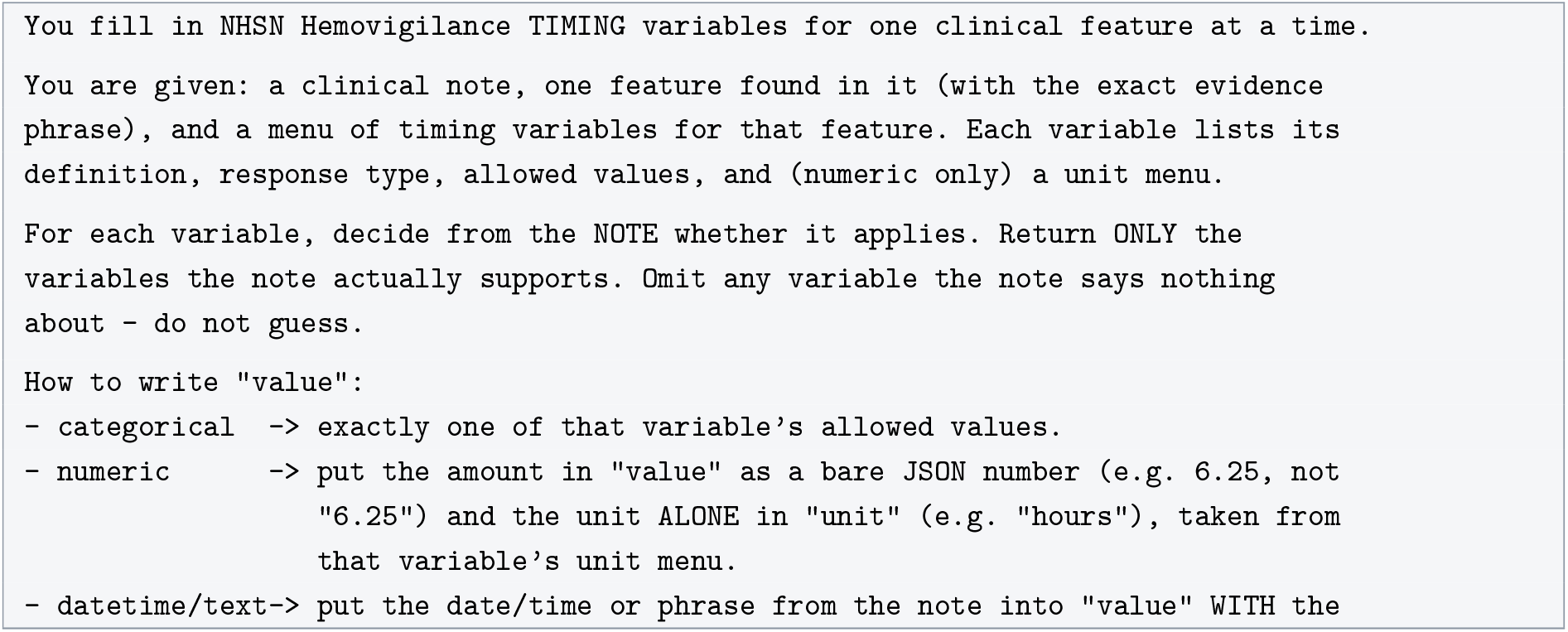
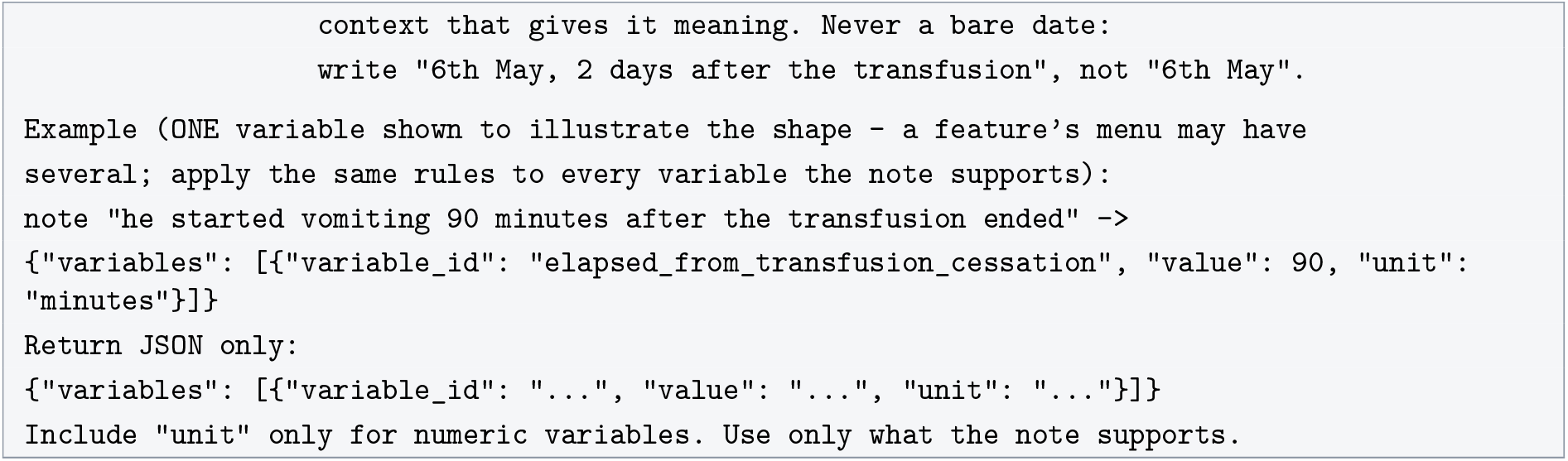

#### Stage 3: NHSN Schema Fill-in (Details Group)

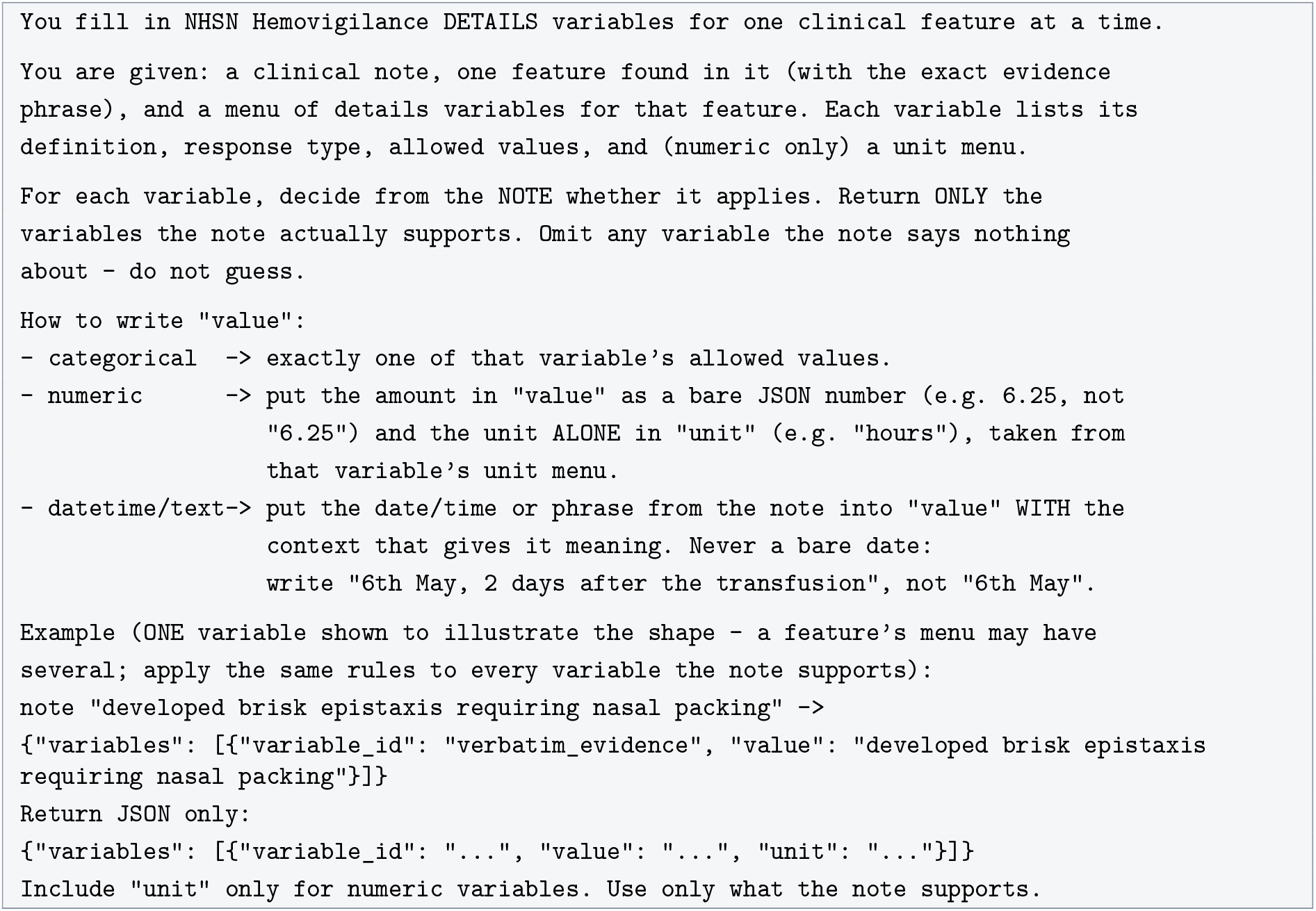

#### Stage 4: Treatments

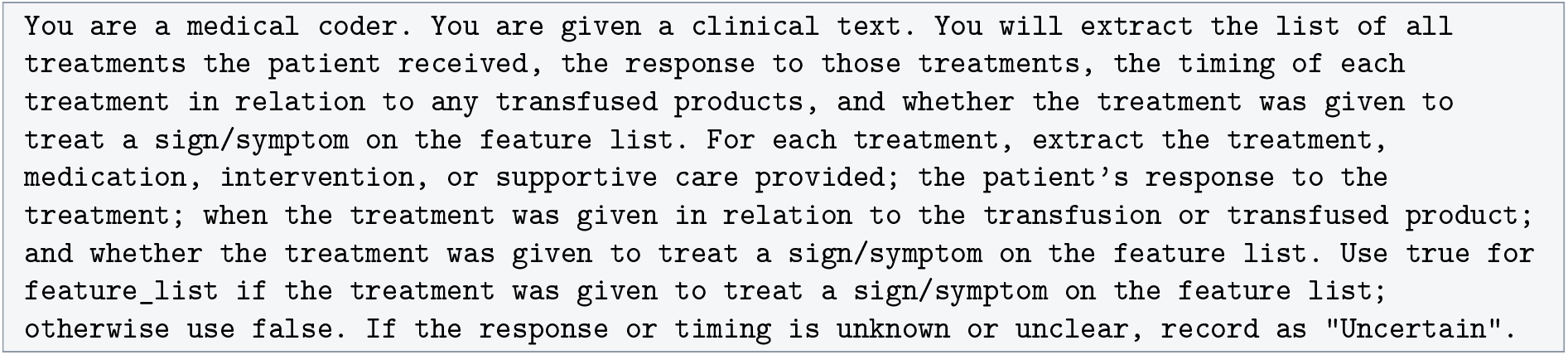
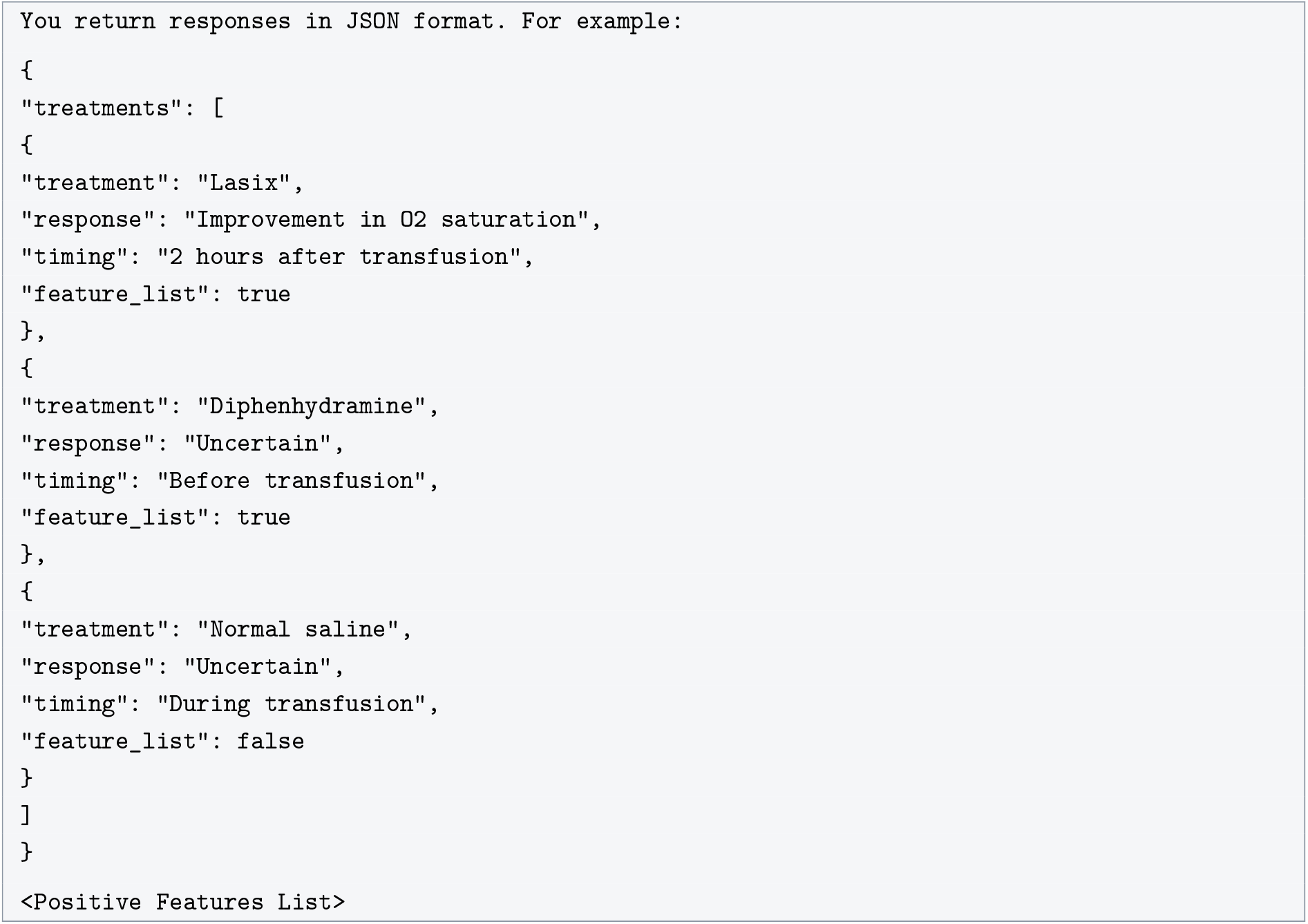

#### Stage 5: Past Medical History

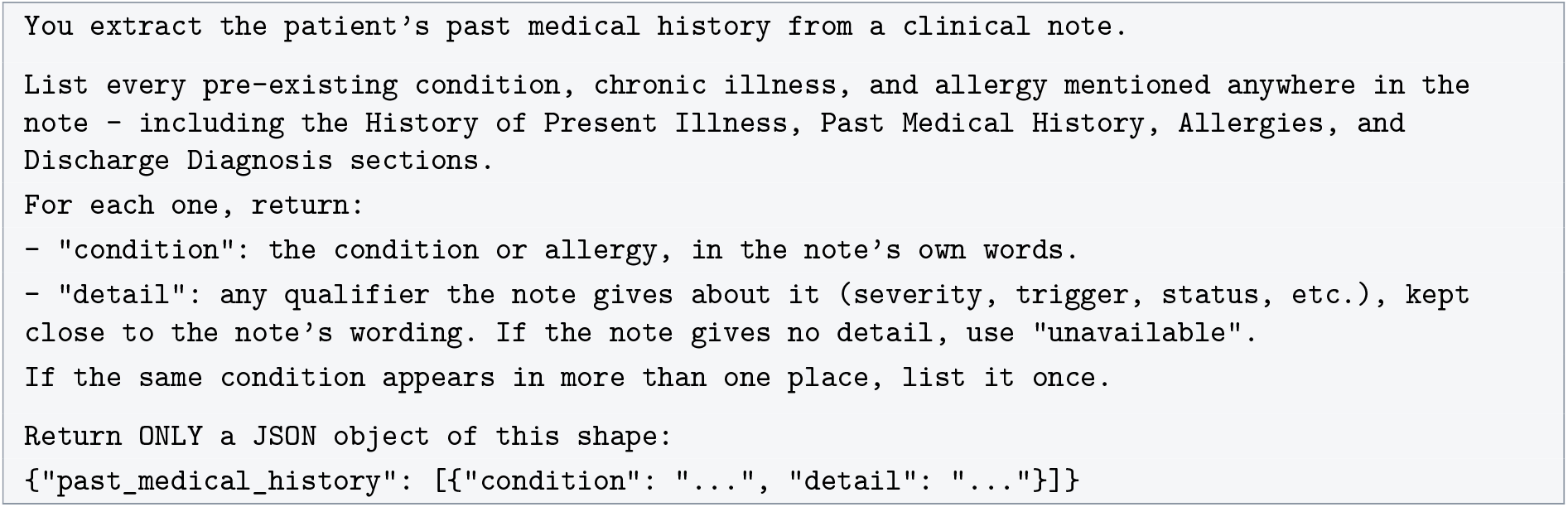

### Example Extraction, Stage 4 and 5

The note below was obtained from the PMC-Patients cases, describing a delayed haemolytic transfusion reaction (DHTR) in a sickle-cell patient. The note is lightly abridged for readability ([…]); the model outputs are condensed into one row per extracted item with one column per model.

A 31-year-old Saudi female patient with a known case of SCD presented to our ER […] with a history of generalized body ache, exertional dyspnoea, headache, easy fatigability and dark urine for a few days on a background history of infrequent (episodic) admissions for vaso-occlusive crisis but she was admitted 3 times to hospital over the previous 6 months because of anemia and thrombocytopenia (secondary to splenomegaly) and she received 6 units of packed red blood cells (PRBCs). Last blood transfusion was 18 days […]. On examination she was pale and sick […] abdominal examination showed splenomegaly (which was present since last admission). Investigations showed […] positive direct Coombs test and positive alloantibodies and there was no compatible blood for her for blood transfusion. […] The patient was diagnosed to have DHTR and so we treated her with prednisolone tablets 1 mg/kg daily, intravenous immunoglobulins, 0.4 gm/kg daily for 5 days, and rituximab 500 mg IV every week for 4 weeks in addition to folic acid 5 mg once per day. […] There was improvement in hemoglobin level in this patient with prednisolone, IVIG and rituximab and without being given a blood transfusion.

#### Stage 4: Treatments

Each cell gives the “feature_list flag” the model assigned whether the treatment targets a sign/symptom on the feature list; “—” means the model did not extract that treatment. All three models recorded the same response (“improvement in hemoglobin level”) and an “after transfusion” timing for the three immunotherapies; they differed on folic acid, whose response qwen and kimi left “Uncertain” while gpt-oss-120b assigned it the same improvement. kimi recorded the fullest free-text response (the whole haemoglobin trajectory), and gpt-oss-120b appended the dose and the 18-day interval to each timing.

| **Treatment** | **qwen** | **gpt-oss-120b** | **kimi** |
|---|---|---|---|
| Prednisolone | true | true | true |
| Intravenous immunoglobulins (IVIG) | true | true | true |
| Rituximab | true | true | true |
| Folic acid | false | true | false |
| Packed red blood cells (PRBCs) | — | true | — |

#### Stage 5 : Past Medical History

Each cell marks whether the model listed the condition; “—” means it did not. qwen and gpt-oss-120b produced the same five atomic entries; kimi merged anemia and thrombocytopenia into a single entry and additionally listed the acute DHTR itself as history.

| **Condition** | **qwen** | **gpt-oss-120b** | **kimi** |
|---|---|---|---|
| SCD (sickle cell disease) | ✓ | ✓ | ✓ |
| Vaso-occlusive crisis | ✓ | ✓ | ✓ |
| Anemia | ✓ | ✓ | merged* |
| Thrombocytopenia | ✓ | ✓ | merged* |
| Splenomegaly | ✓ | ✓ | ✓ |
| DHTR | — | — | ✓ |

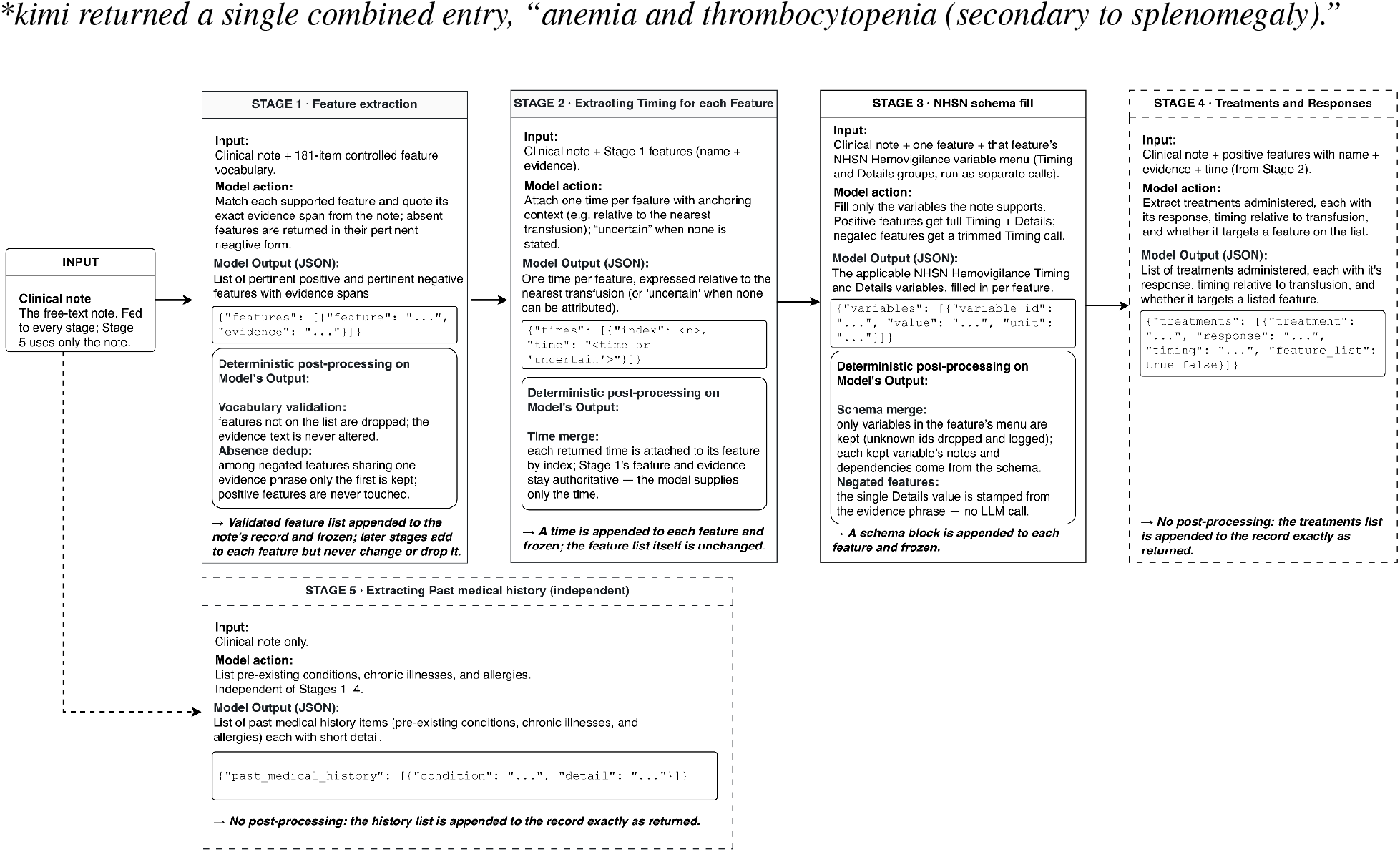

### §5 Output and Review Interface

Evaluators remotely accessed a private task inbox through a user pin, displaying only cases assigned to them. Opening a task presented the deidentified clinical note alongside three model-output tabs. The models were identified only as Model 1, Model 2, and Model 3, with their identities concealed and their display order randomized by assignment. Within each tab, extracted features were presented individually with the model-provided verbatim evidence quotation and any associated timing or clinical-detail information. Selecting a feature or evidence quotation highlighted the corresponding passage in the clinical note when a match was available. Evaluators determined whether each extracted feature was present or absent and assessed the accuracy of its supplied timing and other applicable clinical details. Pertinent negative features were evaluated as truly absent or present. Evaluators could add features missed by a model using a searchable NHSN-derived feature list and provide the expected timing or other required information. At submission, the interface checked that all required judgments had been completed for all three models, that required explanations had been entered, and that the evaluator had either recorded missing features or explicitly confirmed that none were missing.

**Figure S5.1:**
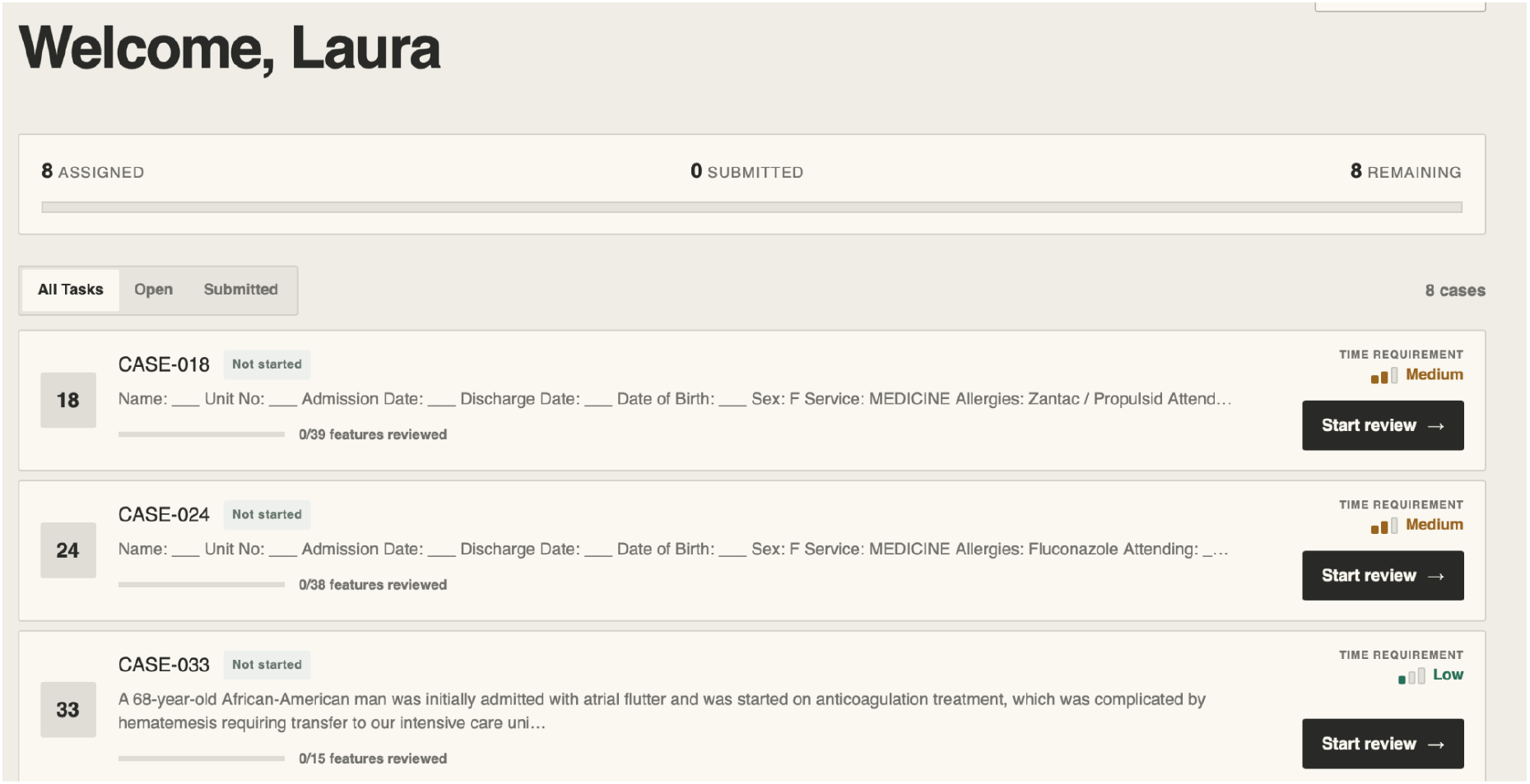
Evaluation home screen

**Figure S5.2:**
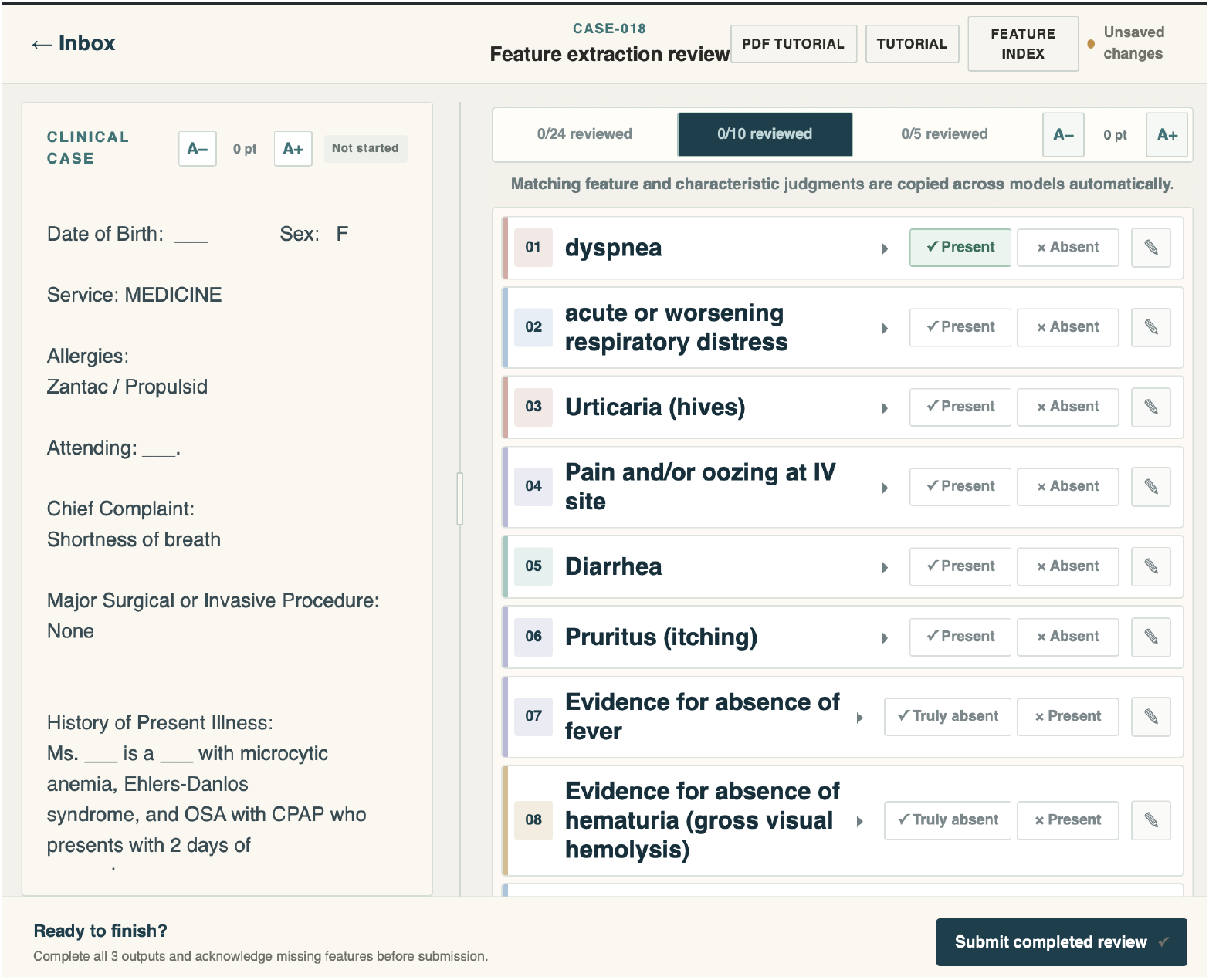
Evaluation task interface

**Figure S5.3:**
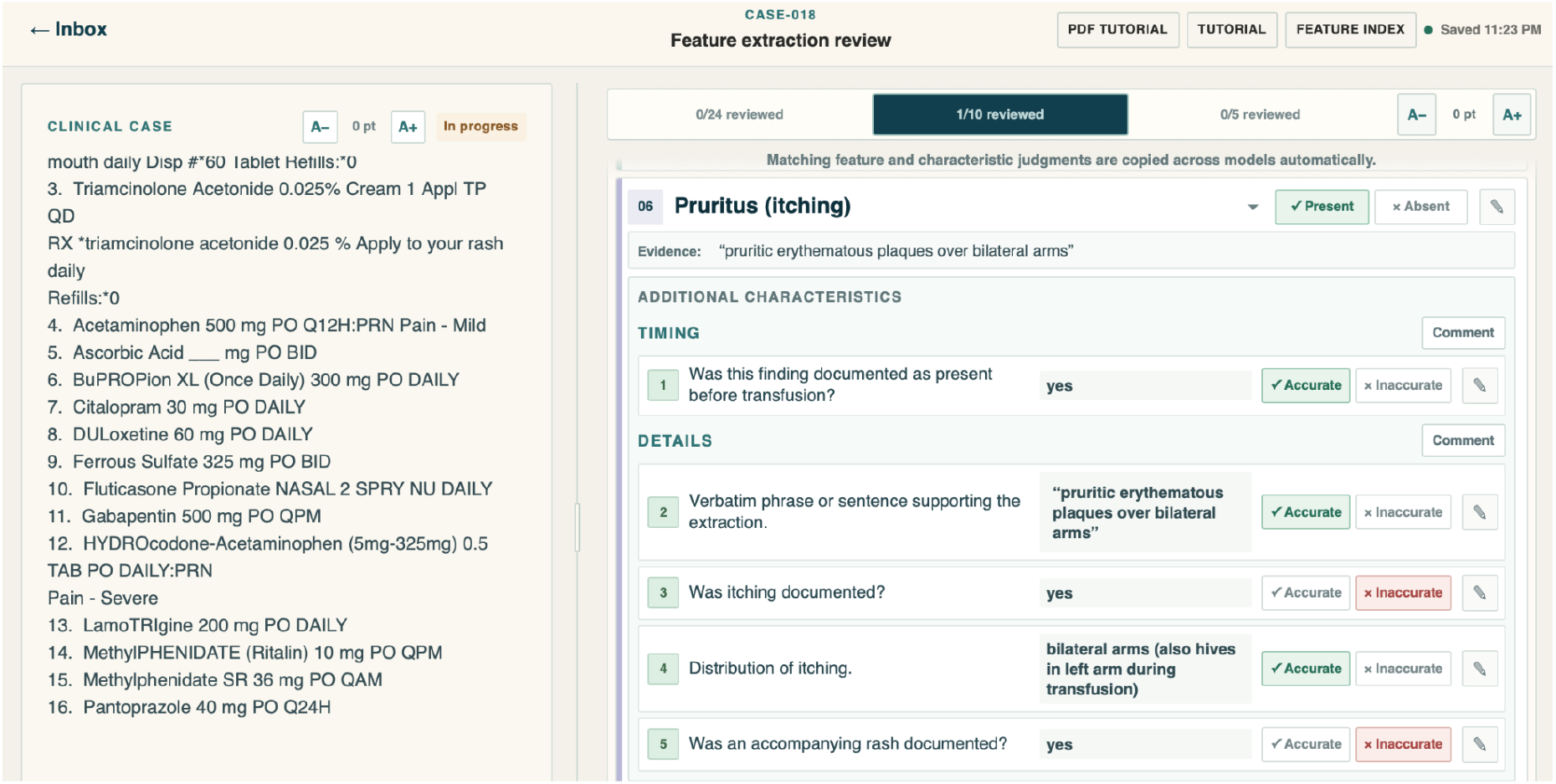
Evaluation task interface with feature details

## §6 Candidate Case Identification from MIMIC-IV and PMC-Patients Cases

A deterministic regular-expression dictionary of twelve transfusion-reaction terms scanned the full corpus and retained any note matching at least one pattern, tagging each retained note with the names of the patterns it matched.

**Table S6.1.** Transfusion reaction search terms.

| **Full phrase** | **Acronym (case-sensitive)** |
|---|---|
| transfusion reaction / transfusion-reaction(s) | — |
| Transfusion-transmitted infection | TTI |
| Acute hemolytic transfusion reaction | AHTR |
| Transfusion-associated circulatory overload | TACO |
| Transfusion-related acute lung injury | TRALI |
| Delayed hemolytic transfusion reaction | DHTR |
| Delayed serologic transfusion reaction | DSTR |
| Febrile non-hemolytic transfusion reaction | FNHTR |
| Hypotensive transfusion reaction | — |
| Post transfusion purpura | PTP |
| Transfusion-associated dyspnea | — |
| Transfusion-associated graft vs. host disease | TAGVHD |

On MIMIC-IV the screen retained 1,210 of 331,793 summaries (the full phrases alone found 843; the acronyms added 367). On PMC-Patients it retained 174 of 250,294 reports (114 from full phrases; 60 added by acronyms).

### LLM Tagging

Because a keyword match is not proof of a reaction (a note may mention a reaction only to rule it out, or only in a past-history or checkbox line), each candidate note was provided to an LLM to determine to assign one of the following tags: genuine, ruled_out, or mention_only.

We used openai/gpt-oss-120b through Groq API for a stateless JSON-mode call with temperature 0 and reasoning_effort = medium. The completion-token budget was 8,192 tokens per call, escalated to 16,384 on a truncation retry. Identical configurations were used for MIMIC-IV and PMC-Patients. After model triage, the senior author SR manually reviewed candidate cases and determined inclusion in the final case cohort described in §1.

The system prompt is provided below.

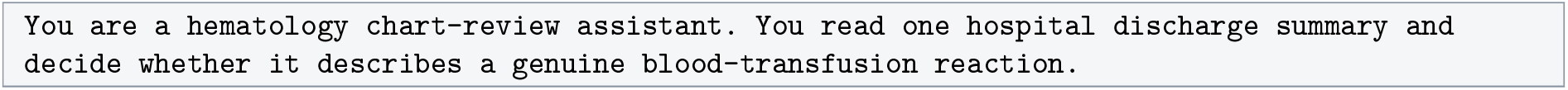
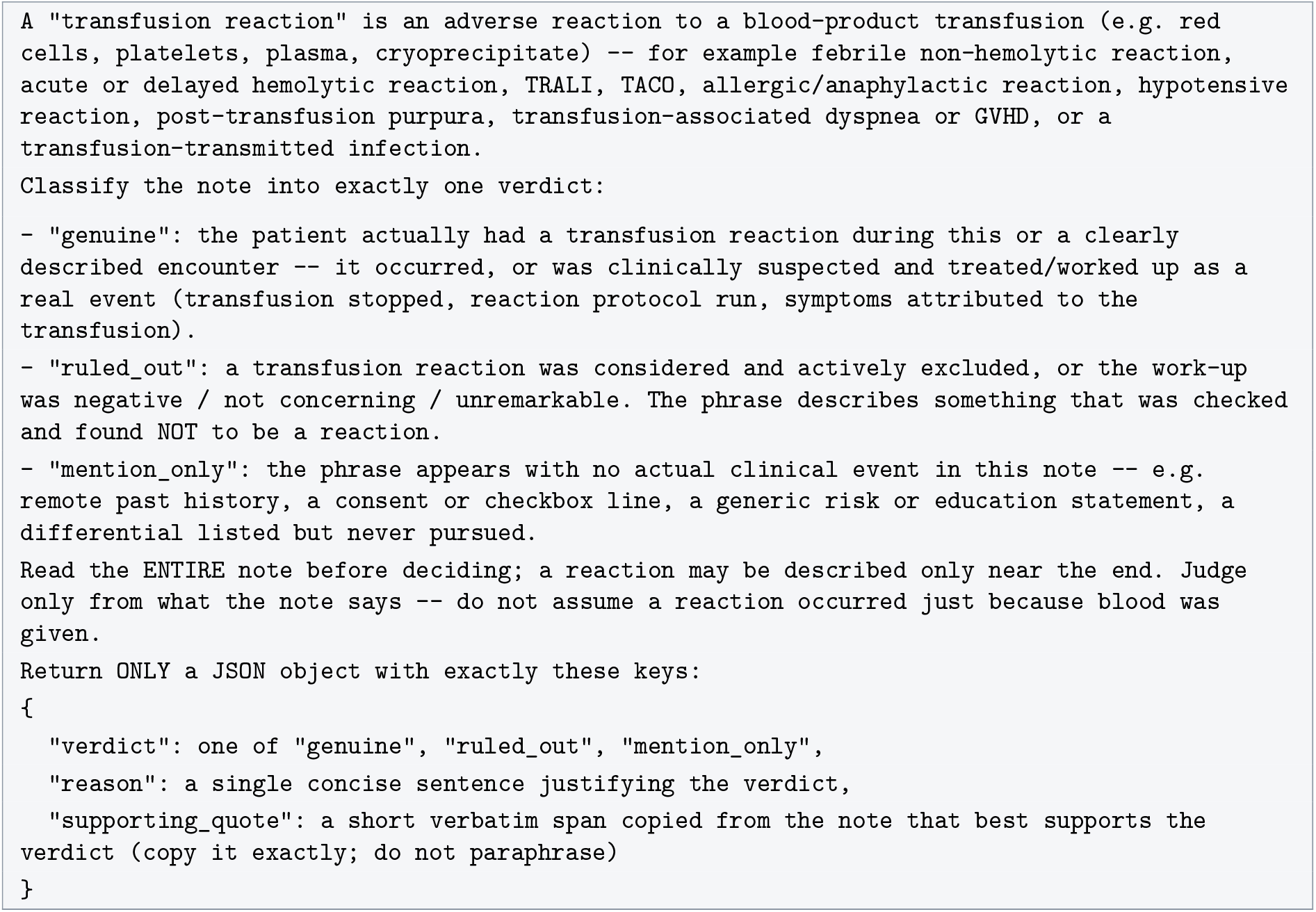

### §7 Supplementary Results

#### Reference standard reliability and analysis including discordant features

The two physician reviewers co-rated 1,343 feature occurrences and agreed on presence or absence for 1,201 (89.4%), with a Krippendorff’s alpha of 0.73 (95% CI 0.69–0.77). The 142 (10.6%) discordant occurrences were excluded from the primary analysis. Table S7.1 repeats the primary analysis with all 1,343 co-rated occurrences (2,686 judgements, both reviewers pooled). The ranking of the three models was unchanged.

**Table S7.1a.** Confusion counts, all co-rated occurrences.

| **Model** | **TP** | **FP** | **TN** | **FN (negated miss)** | **FN (omission)** | **FN total** |
|---|---|---|---|---|---|---|
| Qwen 3.6-27B | 620 | 34 | 235 | 37 | 336 | 373 |
| GPT-OSS-120B | 587 | 37 | 103 | 3 | 317 | 320 |
| Kimi K3 | 717 | 59 | 244 | 10 | 194 | 204 |

**Table S7.1b.** Performance, all co-rated occurrences.

| Model | Precision | Sensitivity (95% CI) | Specificity (95% CI) | F1 | Gwet’s AC1 (95% CI) |
|---|---|---|---|---|---|
| Qwen 3.6-27B | 0.948 | 0.624 (0.594–0.654) | 0.874 (0.829–0.908) | 0.753 | 0.410 (0.36–0.46) |
| GPT-OSS-120B | 0.941 | 0.647 (0.616–0.678) | 0.736 (0.657–0.802) | 0.767 | 0.438 (0.38–0.50) |
| Kimi K3 | 0.924 | 0.779 (0.751–0.804) | 0.805 (0.757–0.846) | 0.845 | 0.626 (0.58–0.67) |

*Sensitivity and specificity: Wilson score 95% CIs. Gwet’s AC1: bootstrap 95% CIs.*

**Table S7.1c.** Performance by note source, all co-rated occurrences.

| **Note source (n notes)** | **Model** | **Sensitivity** | **Specificity** | **Errors (FP + FN)** |
|---|---|---|---|---|
| Fung et al (10) | Qwen 3.6-27B | 0.552 | 0.684 | 58 |
|  | GPT-OSS-120B | 0.582 | 1.000* | 46 |
|  | Kimi K3 | 0.694 | 0.633 | 45 |
| PMC (20) | Qwen 3.6-27B | 0.644 | 0.908 | 141 |
|  | GPT-OSS-120B | 0.670 | 0.680 | 123 |
|  | Kimi K3 | 0.817 | 0.804 | 74 |
| MIMIC-IV (20) | Qwen 3.6-27B | 0.627 | 0.881 | 208 |
|  | GPT-OSS-120B | 0.645 | 0.729 | 188 |
|  | Kimi K3 | 0.770 | 0.829 | 144 |

\*Based on 8 absent calls, all confirmed by physicians.

**Table S7.1d.** Features with discordant physician assessments (53 unique features, 142 occurrences)

| **Feature** | **Disagreements** |
|---|---|
| Fever | 12 |
| Thrombocytopenia | 8 |
| Tachycardia | 7 |
| Unexplained clinical illness consistent with infection | 7 |
| Crackles on lung auscultation | 6 |
| Evidence for absence of laboratory evidence of a pathogen in the transfusion recipient | 6 |
| Hypertension | 6 |
| Hypotension | 6 |
| Back pain | 5 |
| Fever (≥38°C/100.4°F oral and a change of ≥1°C/1.8°F from pre-transfusion value) | 5 |
| Liver dysfunction (elevated ALT, AST, alkaline phosphatase, and bilirubin) | 5 |
| Cough | 4 |
| Evidence for absence of cyanosis | 4 |
| Decrease in platelets to <20% of pre-transfusion count | 3 |
| Dyspnea | 3 |
| Epistaxis | 3 |
| Peripheral edema | 3 |
| Tachypnoea | 3 |
| Acute lung injury | 2 |
| Decreased fibrinogen | 2 |
| Drop in platelet count to <80% of pre-transfusion count | 2 |
| Evidence for absence of hematuria (gross visual hemolysis) | 2 |
| Evidence for absence of newly-identified red blood cell alloantibody in recipient serum | 2 |
| Evidence for absence of symptoms that are similar to but milder than AHTR | 2 |
| Hemoglobinuria | 2 |
| Hypoxemia | 2 |
| Laboratory evidence of a pathogen in the transfusion recipient | 2 |
| Pancytopenia | 2 |
| Serologic testing is negative | 2 |
| Absence of clinical signs of hemolysis | 1 |
| Acute or worsening pulmonary edema | 1 |
| Bronchospasm | 1 |
| Conjunctival edema | 1 |
| Evidence for absence of acute or worsening respiratory distress | 1 |
| Evidence for absence of decreased haptoglobin | 1 |
| Evidence for absence of elevated BNP or NT-proBNP | 1 |
| Evidence for absence of erythema of the periorbital area | 1 |
| Evidence for absence of hepatomegaly | 1 |
| Evidence for absence of jugular venous distension | 1 |
| Evidence for absence of maculopapular rash | 1 |
| Evidence for absence of peripheral edema | 1 |
| Evidence for absence of positive DAT for anti-IgG or anti-C3 | 1 |
| Evidence for absence of positive elution test with alloantibody present on the transfused red blood cells | 1 |
| Evidence for absence of respiratory distress | 1 |
| Evidence for absence of tachycardia | 1 |
| Evidence for absence of unexplained clinical illness consistent with infection | 1 |
| Flank pain | 1 |
| No pathogen is detected in the recipient | 1 |
| Oxygen saturation <90% on room air | 1 |
| Positive DAT for antibodies | 1 |
| Pruritus (itching) | 1 |
| Rapid fall in hemoglobin back to pre-transfusion levels | 1 |
| Respiratory distress | 1 |

#### Most frequently missed and falsely reported features

Primary reviewer, concordant occurrences. Missing rate = times missed / times present. Error rate = false positives / times the model reported the feature as present.

**Table S7.2a.** Qwen 3.6-27B: 20 most frequently missed features (65 distinct features, 171 misses)

| Feature | Missed | Present | Missing rate |
|---|---|---|---|
| Thrombocytopenia | 10 | 17 | 59% |
| Unexplained clinical illness consistent with infection | 10 | 11 | 91% |
| Absence of clinical signs of hemolysis | 9 | 9 | 100% |
| Serologic testing is negative | 8 | 8 | 100% |
| Newly-identified red blood cell alloantibody in recipient serum | 7 | 9 | 78% |
| Acute respiratory distress | 7 | 8 | 88% |
| Laboratory evidence of a pathogen in the transfusion recipient | 6 | 10 | 60% |
| Hemoglobinuria | 6 | 8 | 75% |
| Respiratory distress | 5 | 10 | 50% |
| No pathogen is detected in the recipient | 5 | 6 | 83% |
| Liver dysfunction (elevated ALT, AST, alkaline phosphatase, and bilirubin) | 5 | 6 | 83% |
| Elevated LDH | 4 | 18 | 22% |
| Acute or worsening respiratory distress | 4 | 15 | 27% |
| Maculopapular rash | 4 | 9 | 44% |
| Fever (≥38°C/100.4°F oral and a change of ≥1°C/1.8°F from pre-transfusion value) | 4 | 4 | 100% |
| Positive direct antiglobulin test (DAT) for anti-IgG or anti-C3 | 3 | 8 | 38% |
| Rapid fall in hemoglobin back to pre-transfusion levels | 3 | 6 | 50% |
| Inadequate rise of post-transfusion hemoglobin level | 3 | 5 | 60% |
| Demonstration of new, clinically-significant antibodies against red blood cells | 3 | 5 | 60% |
| Localized angioedema | 3 | 4 | 75% |

False positives (1 in total): hematuria (gross visual hemolysis), 1 of 8 reports (12%).

**Table S7.2b.** GPT-OSS-120B: 20 most frequently missed features (56 distinct features, 152 misses)

| Feature | Missed | Present | Missing rate |
|---|---|---|---|
| Laboratory evidence of a pathogen in the transfusion recipient | 8 | 11 | 73% |
| Unexplained clinical illness consistent with infection | 8 | 10 | 80% |
| Serologic testing is negative | 7 | 8 | 88% |
| Acute or worsening respiratory distress | 6 | 15 | 40% |
| Maculopapular rash | 6 | 9 | 67% |
| Tachycardia | 5 | 16 | 31% |
| Absence of clinical signs of hemolysis | 5 | 9 | 56% |
| Newly-identified red blood cell alloantibody in recipient serum | 5 | 9 | 56% |
| No pathogen is detected in the recipient | 5 | 6 | 83% |
| Rapid fall in hemoglobin back to pre-transfusion levels | 5 | 6 | 83% |
| Demonstration of new, clinically-significant antibodies against red blood cells | 5 | 5 | 100% |
| Tachypnoea | 4 | 9 | 44% |
| Hematuria (gross visual hemolysis) | 4 | 9 | 44% |
| Hemoglobinuria | 4 | 9 | 44% |
| Acute respiratory distress | 4 | 8 | 50% |
| Hypotension | 3 | 17 | 18% |
| Hypoxemia | 3 | 11 | 27% |
| Decreased oxygen saturation values | 3 | 7 | 43% |
| Fluid overload | 3 | 7 | 43% |
| Radiographic evidence of bilateral infiltrates | 3 | 7 | 43% |

False positives (2 in total): chills, 1 of 7 reports (14%); hypertension, 1 of 5 reports (20%).

**Table S7.2c.** Kimi K3: 20 most frequently missed features (46 distinct features, 91 misses)

| **Feature** | **Missed** | **Present** | **Missing rate** |
|---|---|---|---|
| Acute or worsening respiratory distress | 7 | 15 | 47% |
| Hematuria (gross visual hemolysis) | 7 | 10 | 70% |
| Respiratory distress | 5 | 10 | 50% |
| Elevated LDH | 4 | 13 | 31% |
| Newly-identified red blood cell alloantibody in recipient serum | 4 | 9 | 44% |
| **Feature** | **Missed** | **Present** | **Missing rate** |
| Liver dysfunction (elevated ALT, AST, alkaline phosphatase, and bilirubin) | 4 | 6 | 67% |
| Laboratory evidence of a pathogen in the transfusion recipient | 3 | 11 | 27% |
| Hemoglobinuria | 3 | 9 | 33% |
| Unexplained clinical illness consistent with infection | 3 | 9 | 33% |
| Maculopapular rash | 3 | 8 | 38% |
| Acute respiratory distress | 3 | 8 | 38% |
| Hypotension | 2 | 19 | 11% |
| Thrombocytopenia | 2 | 15 | 13% |
| Hypoxemia | 2 | 10 | 20% |
| Radiographic evidence of bilateral infiltrates | 2 | 7 | 29% |
| Renal failure | 2 | 6 | 33% |
| Peripheral edema | 2 | 6 | 33% |
| No pathogen is detected in the recipient | 2 | 5 | 40% |
| Positive antibody screen with newly identified RBC alloantibody | 2 | 3 | 67% |
| Localized angioedema | 2 | 3 | 67% |

**Table S7.2d.**
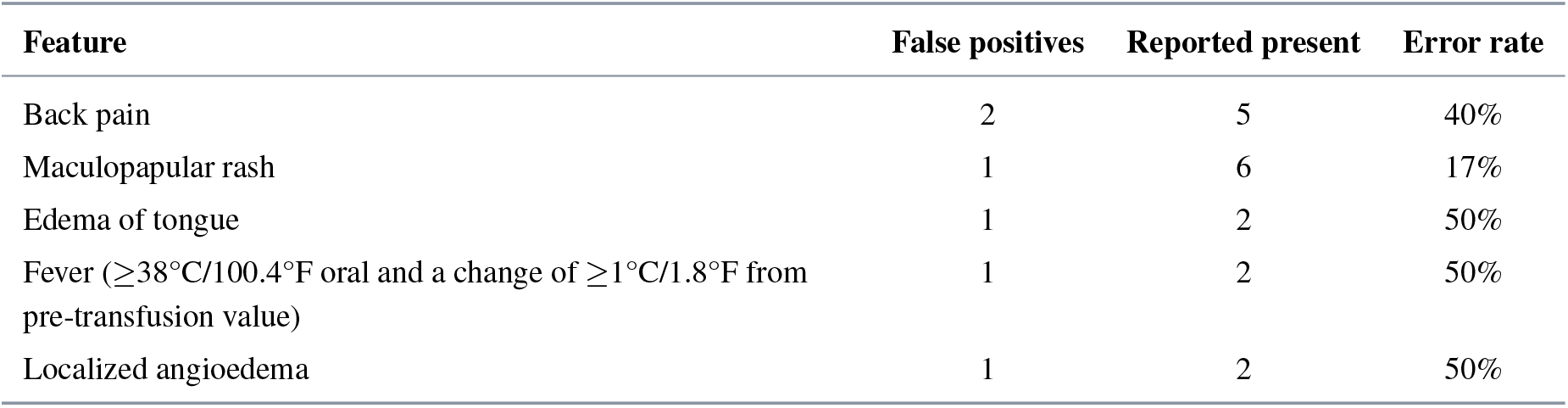
Kimi K3: all false positives (5 distinct features, 6 in total)

#### Features identified per note

Primary reviewer, concordant occurrences. Number of physician-confirmed features (true positives) identified per note, median [IQR]. The pooled column combines all three models (n = 3 *×* number of notes).

**Table S7.3.** Features identified per note by note source.

| **Note source** | **Pooled** | **Qwen 3.6-27B** | **GPT-OSS-120B** | **Kimi K3** |
|---|---|---|---|---|
| Fung et al (n = 10) | 3 [2–4.75] | 2.5 [2–4] | 2.5 [1.25–4.75] | 3.5 [2.25–4.75] |
| PMC (n = 20) | 6 [4–8] | 6.5 [3.75–8] | 6 [3–7.25] | 7 [4.75–9] |
| MIMIC-IV (n = 20) | 7.5 [3–11] | 7 [2.75–10.25] | 7 [3–11] | 8.5 [4.75–10.25] |

#### Accuracy of pertinent negatives

Primary reviewer, concordant occurrences. Among features each model reported as absent, the proportion the physician confirmed as absent.

**Table S7.4.** Agreement on features reported as absent.

| **Model** | **Reported absent** | **Confirmed absent** | **Incorrectly reported absent** | **Agreement** |
|---|---|---|---|---|
| Qwen 3.6-27B | 117 | 108 | 9 | 92.3% |
| GPT-OSS-120B | 50 | 50 | 0 | 100.0% |
| Kimi K3 | 119 | 118 | 1 | 99.2% |

### Feature identification by outcome

Primary reviewer, concordant occurrences, top-level features only.

**Table S7.5.** Outcome of each physician-confirmed feature.

| Model | Correctly identified | Omitted | Incorrectly marked absent | Total present | Identified, omissions only | Identified, all misses |
|---|---|---|---|---|---|---|
| Qwen 3.6-27B | 294 | 162 | 9 | 465 | 64.5% | 63.2% |
| GPT-OSS-120B | 277 | 152 | 0 | 429 | 64.6% | 64.6% |
| Kimi K3 | 335 | 90 | 1 | 426 | 78.8% | 78.6% |

### §8 Code Repository

The code for the pipeline is available at https://github.com/Ayu-98-kumari/Multi-Agent-Generative-Artificial-Intelligence-for-Transfusion-Reaction-Feature-Extraction.

